# Association of Women-Specific Health Factors in the Severity of Parkinson’s Disease

**DOI:** 10.1101/2022.10.03.22280671

**Authors:** Shilpa C Rao, Yadi Li, Brittany Lapin, Sreya Pattipati, Kamalini Ghosh Galvelis, Anna Naito, Nicolas Guitierrez, Thiago Peixoto Leal, Amira Salim, Philippe A. Salles, Maria De Leon, Ignacio F Mata

**Affiliations:** Genomic Medicine Institute, Lerner Research Institute, Cleveland Clinic Foundation, Cleveland, OH; Department of Molecular Medicine, Case Western Reserve University, Cleveland, OH; Department of Quantitative Health Sciences, Lerner Research Institute, Cleveland Clinic Foundation, Cleveland, OH; Center for Outcomes Research and Evaluation, Neurological Institute, Cleveland Clinic Foundation, Cleveland, OH; Parkinson’s Foundation, Miami, FL; Neurological Institute, Cleveland Clinic Foundation, Cleveland, OH; Defeatparkinsons, Nacogdoches, TX

## Abstract

**Background:** Parkinson’s disease (PD) is an age-related neurological disorder known for the observational differences in its risk, progression, and severity between men and women. While estrogen has been considered to be a protective factor in the development of PD, there is little known about the role that fluctuations in hormones and immune responses from sex-specific health experiences have in the disease’s development and severity.

**Objective:** We sought to identify women-specific health experiences associated with PD severity by developing and distributing a women-specific questionnaire across the United States.

**Design:** We created a questionnaire that addresses women’s specific experiences and their PD clinical history and deployed it through The Parkinson’s Foundation: PD Generations. To determine the association between women-specific health factors and PD severity, we constructed multivariable logistic regression models based on the MDS-UPDRS scale and the participants’ questionnaire responses, genetics, and clinical data.

**Results:** For our initial launch in November 2021, we had 304 complete responses from PD GENEration. Univariate and multivariate logistic modeling found significant associations between major depressive disorder, perinatal depression, natural childbirth, *LRRK2* genotype, B12 deficiency, total hysterectomy and increased PD severity.

**Conclusions:** This study is the first nationally available questionnaire for women’s health and PD. It shifts the paradigm in understanding PD etiology and acknowledging how sex-specific experiences may contribute to PD severity. In addition, the work in this study sets the foundation for future research to investigate the reasons behind the sex differences in PD.

## Introduction

As the average age of the global population increases, neurodegenerative diseases, like Parkinson’s disease (PD), are a rising cause of concern because of the consequential global economic and societal burdens. With a drastically increased prevalence in the past two decades, PD is one of the fastest-growing neurological diseases worldwide, affecting ∼1–3% of the population over 60 years old^1,2^. It is characterized by α-synuclein pathology and the loss of dopaminergic neurons in the substantia nigra pars compacta. It is traditionally diagnosed by the presentation of motor abnormalities, like the slowness of movements, rigidity, and tremors. However, other symptoms, such as cognitive dysfunction, depression, and sleep disturbances, can be seen at disease onset as well^3,4^.

PD research has made substantial progress in deciphering what factors influence the disease’s risk, progression, and severity. Genetics, environmental factors, and clinical history all contribute to the etiology of PD. However, due to its multifactorial nature, the root cause of PD is still not known. For the past two decades, highly-penetrant, rare alterations in several genes have been associated with typical familial PD, and common genetic variability at 90 loci has been linked to PD risk, onset, progression, and treatment response^5^. Epidemiological factors associated with PD etiology include diet, occupation, heavy metal/pesticide exposure, and social habits^5,6^. The interactions between all of these factors alongside clinical history (i.e., age, sex, ethnicity, surgeries, medications, and comorbidities) give rise to endless possible combinations that could explain the diverse spectrum of PD subtypes and etiology. As a result, each PD individual experiences a different health-related quality of life, which leads to treatments not being successful across the entire PD population since they have not been able to completely address the diverse spectrum of symptoms.

One important factor in PD etiology is biological sex. Estrogen being a protective factor against PD is a popular thought in the field and seems justified with men being about 1.5 times more likely to have PD than women^7,8^. Aside from risk, PD varies in presentation, severity, and treatment success between sexes^9,10^. For example, women with PD have increased mortality compared to men with PD^11^. Furthermore, one study found that men and women with PD have different rates of progression and severity when adjusted for demographic variables^4^. Regarding PD severity in motor symptoms, two studies showed that there was poorer postural stability in women and greater rigidity in men^12,13^. For the severity of non-motor symptoms, women reported a higher severity of fatigue and mood-related symptoms^14,15^. A few studies also relayed that women with PD tended to have a greater severity of depression and impairment in activities of daily living^12,13^. Thus, while previous studies have described potential sex differences in the epidemiology and clinical expression of PD between men and women, understanding the role of women-specific health factors (WSHFs) is vital because women have been neglected in both research and clinical discussions^16^.

Only a few studies have examined the association between the disease and certain individual WSHFs, such as menses, contraceptives, surgeries, hormonal disorders, pregnancies, hormone-replacement therapy, and menopause. Moreover, the results have been highly contradictory^7,17–19^, which could be possible due to the lack of attention towards these sex-specific factors, as well as the neglect of gathering the necessary data. This study not only includes WSHFs as equally weighted variables in PD severity but also allows for the data to be collected in a comprehensive way through the questionnaire.

Beyond the lack of representation of women’s health in research, WSHFs are often ignored or excluded in clinical trials in and outside the PD field. One study found that ∼42% of clinical trial protocols required contraception or sterility for women without providing any explanation, making women have a lower chance of being included in a clinical trial and resulting in a lower representation of WSHFs in clinical research. Without incorporating these WSHFs, the scientific community ignores the basic biology of women and continues to inaccurately represent the clinical manifestations and treatment outcomes for 50% of the population^20^.

This study seeks to bridge the gaps of knowledge on the role WSHFs play in the severity of PD in women by deploying a national questionnaire and analyzing the responses together with the participants’ genetic and clinical histories. It sets the stage for acknowledging sex-specific experiences in an age-related neurological disorder and will inform both women and clinicians about what WSHFs contribute to PD severity.

## Methods

### Questionnaire Development and Implementation

With the gap in information regarding WSHFs and PD, we developed and implemented a nationally distributed questionnaire, as large datasets of patient-reported outcomes within the epidemiological field are often generated by questionnaires^21^. S.C.R and I.F.M compiled a list of WSHFs based on their possible association with the disease or their lack of inclusion in PD data thus far. Neurologists and movement disorder specialists at Cleveland Clinic (H.F. and P.S.), along with PD and women’s health expert (M.D.L), advised on further modifications of the questionnaire. It was then formatted using a combination of short-text (age and additional comments to be typed if needed), checkboxes, and Likert-type rating scales for symptom changes with the guidance of the Cleveland Clinic Quantitative Health Sciences patient-reported outcomes team (B.L.). Our primary concern was to limit recall bias, as our subjects skewed towards being above the age of 50 and post-menopausal, so we implemented branching logic, allowing the participant to answer questions that are relevant based on their age of diagnosis (during menses, before or after menopause) and women-specific experiences (including birth control, pregnancies, surgeries, and hormone-replacement therapy) that can occur around that age range. The questionnaire, which can be found in its entirety in our previous work^22^, is on REDCap, a secure web application used for building online surveys and databases^23^. To validate that our questionnaire was comprehensible and could be completed in a reasonable time, movement disorder specialists at CCF administered it with a tablet in the waiting room to 10 women with PD and verified that these subjects were able to answer the questions comprehensively in a timely manner. The participants took an average of 20 minutes to complete the questionnaire.

The Parkinson’s Foundation launched the PD GENEration study (PD GENE; ClinicalTrials.gov identifier: NCT04057794), which provides genetic testing for seven PD-related genes by a CLIA-certified laboratory and genetic counseling at multiple locations in North America at no cost to the participant. At the same time, they collect critical clinical and demographic information from each patient^24^. Each participant is seen and clinically assessed by movement disorder specialist, who inputs their demographic and clinical data. We have partnered with the Parkinson’s Foundation to implement our questionnaire because, in the past, PD GENEration has successfully distributed various research questionnaires from other institutions to patients on their email list who consented to receive questionnaires designed for research^25,26^. In mid-November 2021, PD GENEration released our questionnaire to PD GENEration women.

PD GENEration has IRB approval (IRB # 20-596) to share de-identified patient data (genetics, clinical information, and demographics) in addition to the responses to our questionnaire with us for research purposes. PD GENEration exported the participants’ responses alongside their genetic and clinical data de-identified. This study is approved by Cleveland Clinic (IRB # 21-1138).

### Quality Control

As this is a patient-reported study, we filtered the variables in the questionnaire and demographic data from PD generation, we removed variables where at least 80% of responses are missing or “not available”.

### MDS-UPDRS Severity Scale

The Unified Parkinson’s Disease Rating Scale (MDS-UPDRS) is the most widely used clinical scale for rating motor and non-motor symptom severity in PD. The MDS-UPDRS has four parts: I: Non-motor Experiences of Daily Living; II: Motor Experiences of Daily Living; III: Motor Examination; IV: Motor Complications^27^. At the time of recruitment, PD GENEration had either a clinician or study coordinator administer the MDS-UPDRS.

Missing data in clinical rating scales is problematic because assessments are usually time-locked to an office visit and cannot be retrospectively filled in with reliability. To overcome the missingness in the dataset, we followed the protocol set by Goetz et al. that established thresholds for the maximum number of scores that can be missing from each part. Goetz et al. found that only one missing item from Part I, two from Part II, seven from Part III, and zero from Part IV can be allowed for an individual to be included in subsequent analyses^28^. If the number of missing items is within the threshold, we followed a protocol implemented by Goetz et al. to create a prorated total score for UPDRS Parts I-IV^28^.

Following these adjustments, we divided our cohort into “mild” and “moderate or severe” UPDRS groups based on the maximum triangulation cut-off values stated in Martinez-Martin et al ^29^. We decided to group them by “mild” vs. “moderate/severe” due to the low number of women with a “severe” UPDRS score. Thresholds for moderate/severe were Part I: >21; Part II: >29; Part III: >58; and Part IV: >12. Mild vs. moderate/severe distributions for each UPDRS part are presented in **Supplemental Table 1**.

### Statistical analysis

Demographics, clinical characteristics, genetics, questionnaire responses, and UPDRS scores were summarized using descriptive statistics. Univariate logistic regression models were constructed for each UPDRS subpart to evaluate the associations between PD severity and WSHFs, genetics, and clinical variables. For “check all that apply” questions, dummy variables were created for each response option. Variables with fewer than five positive responses were not included in the analysis. Furthermore, for Likert type questions, “N/A” and “Don’t recall” were set to missing. Questions were excluded if more than 80% of responses were “N/A” or “Don’t recall.” As our dataset contains novel variables that have neither been explored nor been well understood in the PD field so far, we first sought to see which variables may be associated with predicting PD severity using a univariate logistic regression model. If a variable had a very low or high prevalence, the Firth method was used in our regression modeling^30,31^. For each UPDRS subpart, multivariable logistic regression models were constructed from the variables of interest that were gathered from the univariate regression models. These included age, disease duration, and medication, as determined a priori, along with WSHF variables that were significant in the univariate logistic regression models at p<0.05. Model assumptions and multicollinearity were assessed. All statistical analyses were conducted using SAS version 9.4 (SAS Institute Inc, Cary, NC). Statistical significance was established throughout at p<0.05.

## Results

### PD GENEration population

The questionnaire was sent via email on November 27, 2021 to 966 women with PD enrolled with the Parkinson’s Foundation PD GENEration. An email reminder was sent in mid-December, and the questionnaire closed on December 27, 2021. Of the returned surveys, 304 women gave complete responses, providing a response rate of 31.5%. Overall, respondents averaged 64.7 years in age (±9.1), with a mean age at diagnosis of 58.5 years (±10.4). Women completing the survey had an average of 6.2 years of disease duration (±5.3) (since they were diagnosed with PD) (**Table 1**). Since PD GENEration tests participants for seven known PD genes ^24,32^, we are able to see the genetic status of each respondent. Within our cohort, we have women with PD with *GBA* (10%), *LRRK2* (8%), *PINK1* (1%), *PRKN* (4%), and *VPS35* (1.3%) mutations. The entirety of demographic information collected for every PD GENEration respondent by severity group can be found in **Table 1**.

**Table 1A.** Characteristics of the Study Sample: Stratified by Part I Severity Groups.

|  | Total | Mild | Moderate /Severe | p-value |
| --- | --- | --- | --- | --- |
| **Sample Size** | 160 | 112 | 48 |  |
| **Age, mean (sd)** | 64.4 (9.2) | 64.1 (9.2) | 65.0 (9.2) | 0.60<sup>a1</sup> |
| **Age at diagnosis, mean (sd)** | 58.2 (10.3) | 58.6 (9.9) | 57.3 (11.3) | 0.45<sup>a1</sup> |
| **Years since symptom onset, mean (sd)** | 8.6 (6.1) | 7.9 (5.7) | 10.4 (6.7) | **0.017<sup>a1</sup>** |
| **Years since diagnosis, mean (sd)** | 6.1 (5.3) | 5.5 (4.7) | 7.7 (6.2) | **0.032<sup>a2</sup>** |
| **Ethnicity, n (%)** |  |  |  | 0.55<sup>d</sup> |
| Hispanic/Latino | 3 (1.9) | 3 (2.7) | 0 (0.00) |  |
| Not Hispanic or Latino | 157 (98.1) | 109 (97.3) | 48 (100.0) |  |
| **Race, n (%)** |  |  |  | 0.89<sup>d</sup> |
| Asian | 1 (0.63) | 1 (0.89) | 0 (0.00) |  |
| Asian, Native Hawaiian or Other Pacific Islander | 1 (0.63) | 1 (0.89) | 0 (0.00) |  |
| Asian, White | 1 (0.63) | 1 (0.89) | 0 (0.00) |  |
| Black or African American | 1 (0.63) | 1 (0.89) | 0 (0.00) |  |
| Black or African American, White | 1 (0.63) | 0 (0.00) | 1 (2.1) |  |
| White | 154 (96.3) | 107 (95.5) | 47 (97.9) |  |
| White, Native American | 1 (0.63) | 1 (0.89) | 0 (0.00) |  |
| **Genetic Status, n (%)\*** |  |  |  | 0.37<sup>d</sup> |
| GBA | 17 (10.8) | 13 (11.8) | 4 (8.3) |  |
| LRRK2 | 13 (8.2) | 8 (7.3) | 5 (10.4) |  |
| PINK1 | 1 (0.63) | 1 (0.91) | 0 (0.00) |  |
| PRKN | 6 (3.8) | 2 (1.8) | 4 (8.3) |  |
| VPS35 | 1 (0.63) | 1 (0.91) | 0 (0.00) |  |
| None | 120 (75.9) | 85 (77.3) | 35 (72.9) |  |
| **Medications, n(%)** |  |  |  |  |
| Levodopa | 116 (72.5) | 78 (69.6) | 38 (79.2) | 0.22<sup>c</sup> |
| Entacapone | 3 (1.9) | 0 (0.00) | 3 (6.3) | **0.026<sup>d</sup>** |
| Carbidopa, Levodopa, and Entacapone (Stalevo) | 12 (7.6) | 6 (5.4) | 6 (12.8) | 0.18<sup>d</sup> |
| Tolcapone (excluding Stalevo) | 0 (0.00) | 0 (0.00) | 0 (0.00) |  |
| Apomorphine (injection, infusion, other) | 0 (0.00) | 0 (0.00) | 0 (0.00) |  |
| Pramipexole | 22 (13.9) | 14 (12.6) | 8 (17.0) | 0.46<sup>c</sup> |
| Ropinirole | 11 (7.0) | 4 (3.6) | 7 (14.9) | **0.017<sup>d</sup>** |
| Rotigotine | 6 (3.8) | 4 (3.6) | 2 (4.3) | 0.99<sup>d</sup> |
| Rasagiline | 33 (20.9) | 25 (22.5) | 8 (17.0) | 0.44<sup>c</sup> |
| Selegiline (oral, sublingual) | 3 (1.9) | 2 (1.8) | 1 (2.1) | 0.99<sup>d</sup> |
| Amantadine (liquid, infusion) | 17 (10.8) | 14 (12.6) | 3 (6.4) | 0.25<sup>c</sup> |
| **Any_medication, n (%)** |  |  |  |  |
|  | 137 (85.6) | 92 (82.1) | 45 (93.8) | 0.055<sup>c</sup> |
| **Current Comorbidities, n (%)** |  |  |  |  |
| ALS | 0 (0.00) | 0 (0.00) | 0 (0.00) |  |
| Alzheimer's disease | 0 (0.00) | 0 (0.00) | 0 (0.00) |  |
| Arrhythmia/Atrial Fibrillation | 10 (6.3) | 7 (6.3) | 3 (6.3) | 0.99<sup>d</sup> |
| Arthritis | 85 (53.1) | 54 (48.2) | 31 (64.6) | 0.057<sup>c</sup> |
| B12 deficiency | 16 (10.0) | 9 (8.0) | 7 (14.6) | 0.25<sup>d</sup> |
| Cancer | 6 (3.8) | 4 (3.6) | 2 (4.2) | 0.99<sup>d</sup> |
| Congestive Heart Failure | 1 (0.63) | 0 (0.00) | 1 (2.1) | 0.30<sup>d</sup> |
| Crohn's disease/ulcerative colitis | 1 (0.63) | 0 (0.00) | 1 (2.1) | 0.30<sup>d</sup> |
| Depression | 44 (27.5) | 18 (16.1) | 26 (54.2) | **<0.001<sup>c</sup>** |
| Diabetes Mellitus (adult onset) | 6 (3.8) | 3 (2.7) | 3 (6.3) | 0.37<sup>d</sup> |
| Diabetes Mellitus (childhood onset) | 1 (0.63) | 1 (0.89) | 0 (0.00) | 0.99<sup>d</sup> |
| Gaucher disease | 1 (0.63) | 1 (0.89) | 0 (0.00) | 0.99<sup>d</sup> |
| Gout | 2 (1.3) | 2 (1.8) | 0 (0.00) | 0.99<sup>d</sup> |
| Hearing Loss | 39 (24.4) | 23 (20.5) | 16 (33.3) | 0.084<sup>c</sup> |
| Hip Fracture | 0 (0.00) | 0 (0.00) | 0 (0.00) |  |
| Hyper cholesterolemia (High Cholesterol) | 44 (27.5) | 28 (25.0) | 16 (33.3) | 0.28<sup>c</sup> |
| Hypertension (High Blood Pressure) | 30 (18.8) | 22 (19.6) | 8 (16.7) | 0.66<sup>c</sup> |
| Liver | 2 (1.3) | 2 (1.8) | 0 (0.00) | 0.99<sup>d</sup> |
| Loss of smell | 68 (42.5) | 49 (43.8) | 19 (39.6) | 0.63<sup>c</sup> |
| Lung disease (including emphysema) | 11 (6.9) | 6 (5.4) | 5 (10.4) | 0.31<sup>d</sup> |
| Multiple sclerosis | 0 (0.00) | 0 (0.00) | 0 (0.00) |  |
| Myocardial Infarction (Heart Attack) | 0 (0.00) | 0 (0.00) | 0 (0.00) |  |
| Normal pressure hydrocephalus | 0 (0.00) | 0 (0.00) | 0 (0.00) |  |
| Other | 60 (37.5) | 36 (32.1) | 24 (50.0) | **0.033<sup>c</sup>** |
| Peptic ulcer disease | 1 (0.63) | 0 (0.00) | 1 (2.1) | 0.30<sup>d</sup> |
| Peripheral neuropathy | 17 (10.6) | 13 (11.6) | 4 (8.3) | 0.54<sup>c</sup> |
| Peripheral vascular disease | 2 (1.3) | 1 (0.89) | 1 (2.1) | 0.51<sup>d</sup> |
| PTSD | 9 (5.6) | 3 (2.7) | 6 (12.5) | **0.022<sup>d</sup>** |
| Recreational drug use | 9 (5.6) | 6 (5.4) | 3 (6.3) | 0.99<sup>d</sup> |
| Renal insufficiency (kidney disease) | 4 (2.5) | 2 (1.8) | 2 (4.2) | 0.58<sup>d</sup> |
| Seizure, fit, convulsion or unexplained loss of consciousness | 2 (1.3) | 0 (0.00) | 2 (4.2) | 0.089<sup>d</sup> |
| Stroke, mini stroke, CVA (cerebrovascular accident) or TIA (transient ischemic attack) | 2 (1.3) | 0 (0.00) | 2 (4.2) | 0.089<sup>d</sup> |
| Thyroid disease (not cancer, including Grave's disease) | 32 (20.0) | 20 (17.9) | 12 (25.0) | 0.30<sup>c</sup> |
| **UPDRS Score** |  |  |  |  |
| Part 1, mean (sd) | 8.5 (6.1) | 5.3 (2.7) | 16.0 (5.1) | **<0.001<sup>a2</sup>** |
| Part 2, mean (sd) | 7.6 (6.7) | 5.6 (4.6) | 12.2 (8.3) | **<0.001<sup>a2</sup>** |
| Part 3, mean (sd)* | 21.4 (12.0) | 20.6 (10.8) | 23.4 (14.2) | 0.26<sup>a2</sup> |
| Part 4, mean (sd)* | 3.0 (3.4) | 2.5 (2.9) | 4.1 (3.9) | **0.012<sup>a2</sup>** |

\*Data not available for all subjects. Missing values: Genetic Status = 2; Entacapone = 1; Carbidopa, Levodopa, and Entacapone (Stalevo) = 2; Tolcapone (excluding Stalevo) = 2; Apomorphine (injection, infusion, other) = 2; Pramipexole = 2; Ropinirole = 2; Rotigotine = 2; Rasagiline = 2; Selegiline (oral, sublingual) = 1; Amantadine (liquid, infusion) = 2; Part 3 = 19; Part 4 = 21. Statistics presented as Mean (SD), N (column %). p-values: <sup>a1</sup>=t-test, <sup>a2</sup>=Satterthwaite t-test, <sup>c</sup>=Pearson's chi-square test, <sup>d</sup>=Fisher's Exact test.

**Table 1B.**
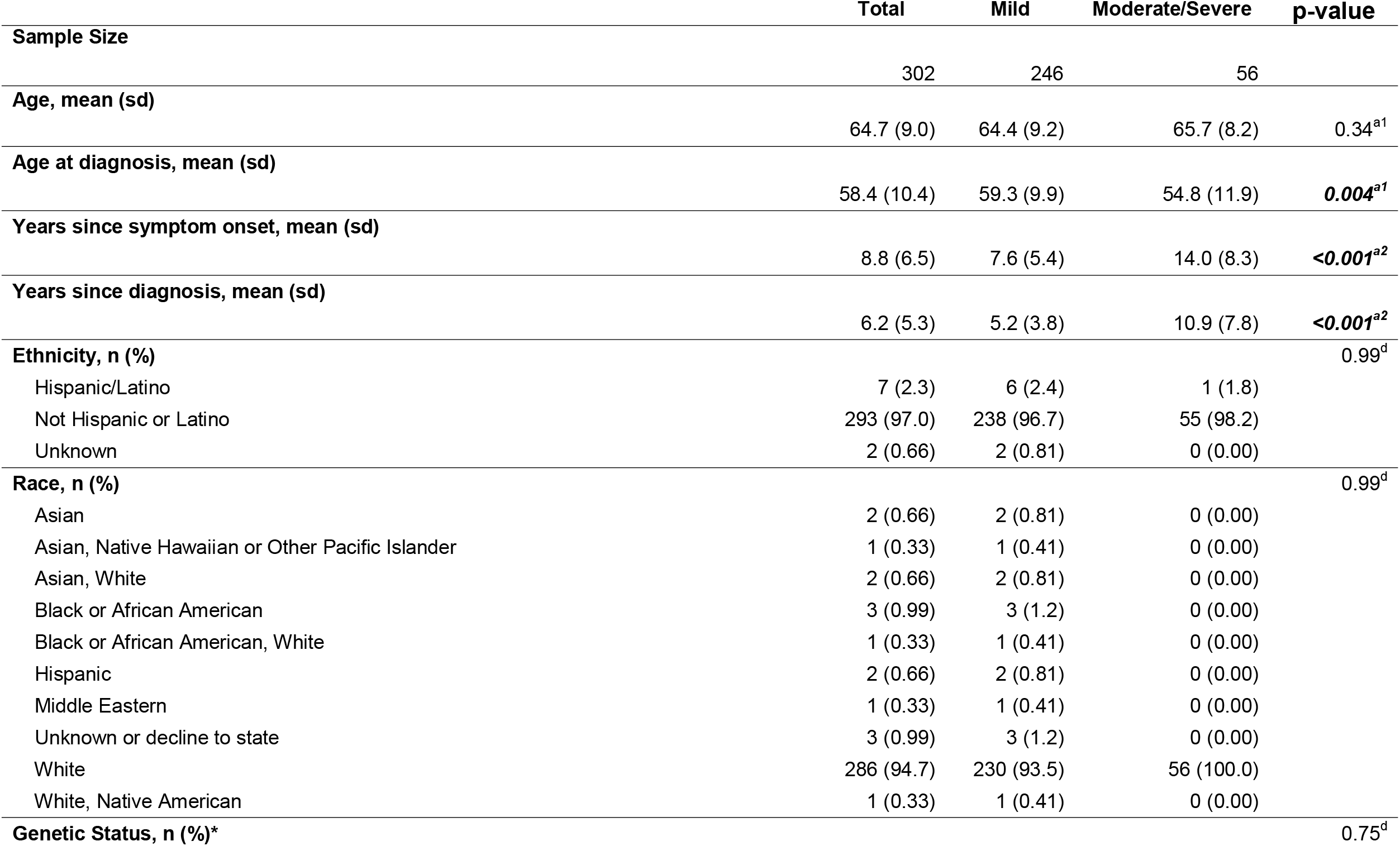
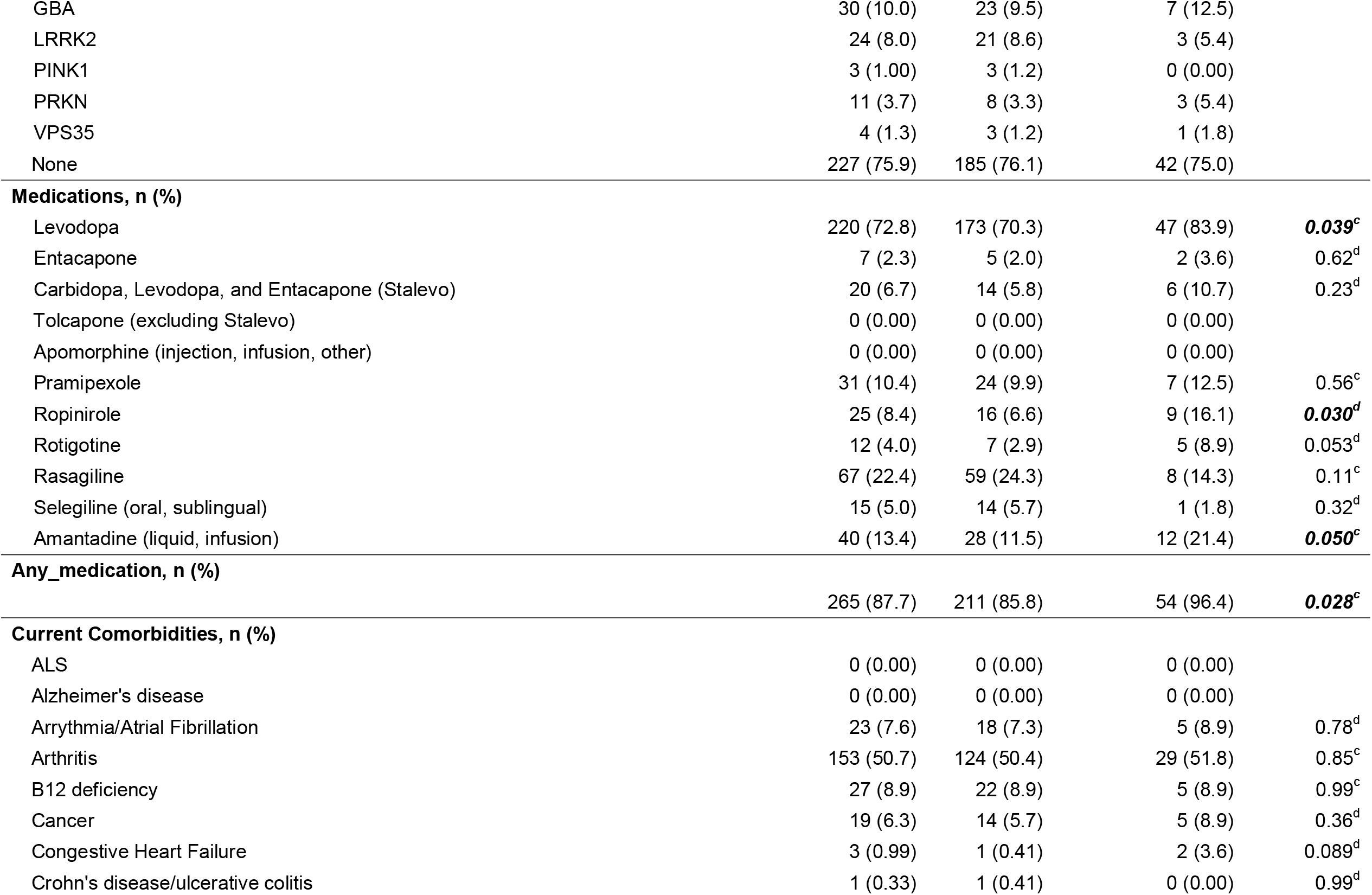
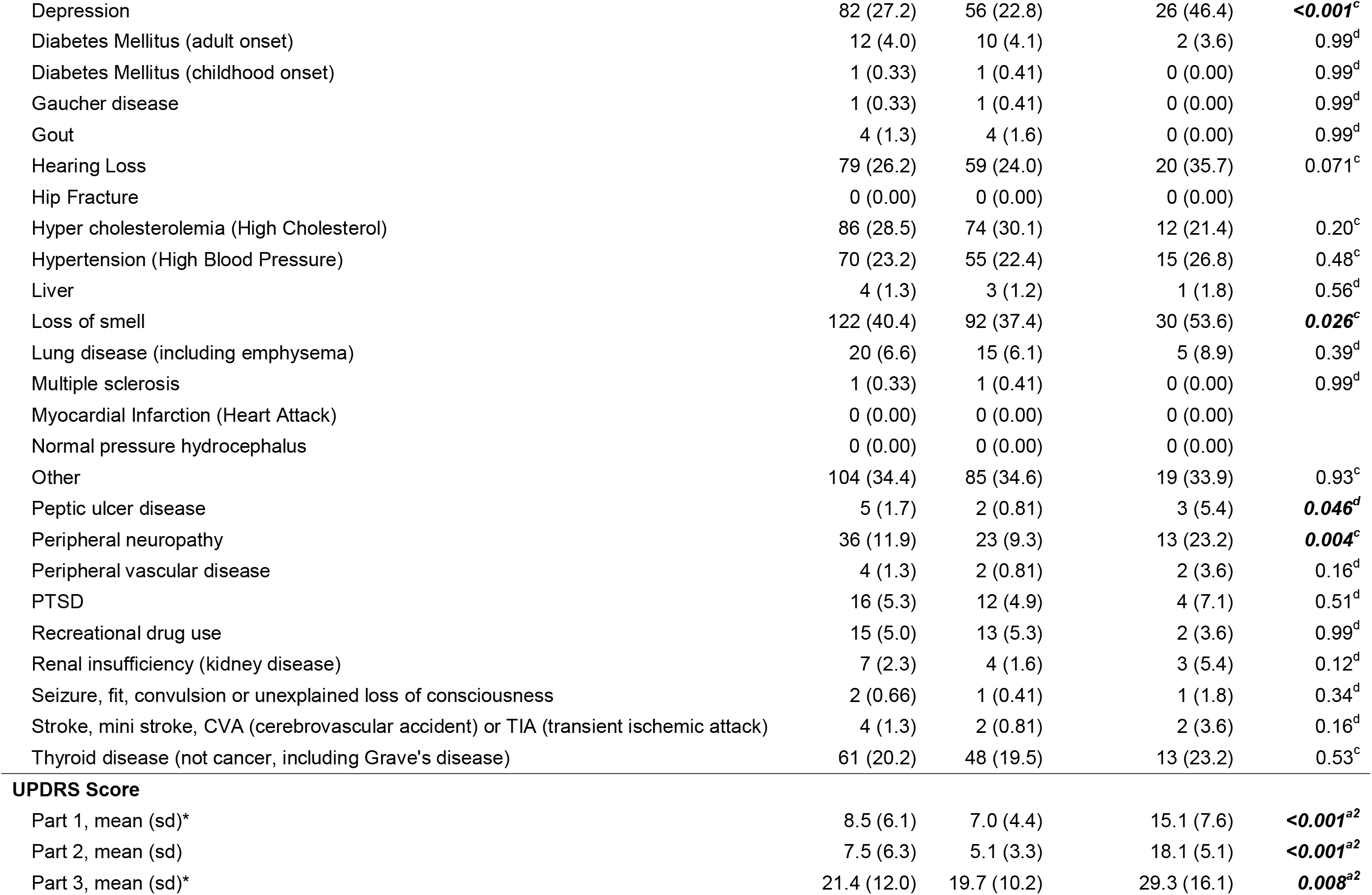
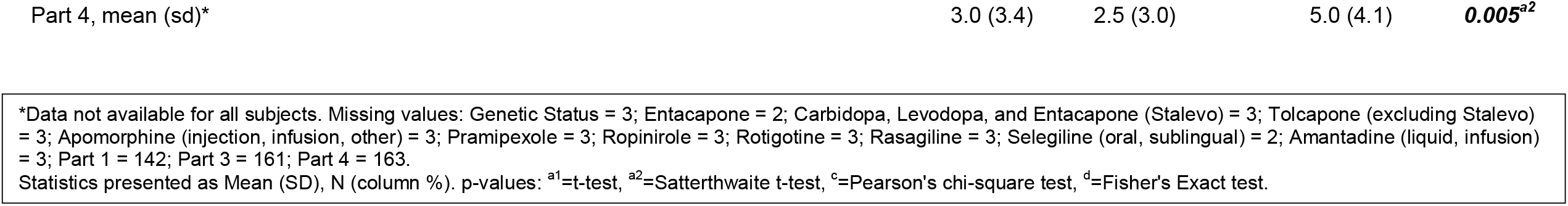
Characteristics of the Study Sample: Stratified by Part II Severity Groups.

**Table 1C.** Characteristics of the Study Sample: Stratified by Part III Severity Groups.

|  | **Total** | **Mild** | **Moderate/Severe** | **p-value** |
| --- | --- | --- | --- | --- |
| **Sample Size** | 141 | 114 | 27 |  |
| **Age, mean (sd)** | 64.1 (8.7) | 64.0 (8.8) | 64.6 (8.5) | 0.72<sup>a1</sup> |
| **Age at diagnosis, mean (sd)** | 57.9 (9.7) | 58.1 (9.5) | 56.9 (10.6) | 0.55<sup>a1</sup> |
| **Years since symptom onset, mean (sd)** | 8.8 (6.0) | 8.5 (5.6) | 10.1 (7.2) | 0.19<sup>a1</sup> |
| **Years since diagnosis, mean (sd)** | 6.2 (5.2) | 5.9 (4.6) | 7.8 (7.2) | 0.19<sup>a2</sup> |
| **Ethnicity, n (%)** |  |  |  | 0.99<sup>d</sup> |
| Hispanic/Latino | 3 (2.1) | 3 (2.6) | 0 (0.00) |  |
| Not Hispanic or Latino | 138 (97.9) | 111 (97.4) | 27 (100.0) |  |
| **Race, n (%)** |  |  |  | 0.58<sup>d</sup> |
| Asian, Native Hawaiian or Other Pacific Islander | 1 (0.71) | 1 (0.88) | 0 (0.00) |  |
| Asian, White | 1 (0.71) | 0 (0.00) | 1 (3.7) |  |
| Black or African American, White | 1 (0.71) | 1 (0.88) | 0 (0.00) |  |
| White | 137 (97.2) | 111 (97.4) | 26 (96.3) |  |
| White, Native American | 1 (0.71) | 1 (0.88) | 0 (0.00) |  |
| **Genetic Status, n (%)\*** |  |  |  | 0.096<sup>d</sup> |
| GBA | 14 (10.1) | 10 (8.9) | 4 (14.8) |  |
| LRRK2 | 11 (7.9) | 6 (5.4) | 5 (18.5) |  |
| PINK1 | 1 (0.72) | 1 (0.89) | 0 (0.00) |  |
| PRKN | 6 (4.3) | 4 (3.6) | 2 (7.4) |  |
| VPS35 | 1 (0.72) | 1 (0.89) | 0 (0.00) |  |
| None | 106 (76.3) | 90 (80.4) | 16 (59.3) |  |
| **Medications, n (%)** |  |  |  |  |
| Levodopa | 103 (73.0) | 85 (74.6) | 18 (66.7) | 0.41<sup>c</sup> |
| Entacapone | 3 (2.1) | 2 (1.8) | 1 (3.7) | 0.48<sup>d</sup> |
| Carbidopa, Levodopa, and Entacapone (Stalevo) | 10 (7.2) | 8 (7.1) | 2 (7.4) | 0.99<sup>d</sup> |
| Tolcapone (excluding Stalevo) | 0 (0.00) | 0 (0.00) | 0 (0.00) |  |
| Apomorphine (injection, infusion, other) | 0 (0.00) | 0 (0.00) | 0 (0.00) |  |
| Pramipexole | 19 (13.7) | 14 (12.5) | 5 (18.5) | 0.53<sup>d</sup> |
| Ropinirole | 10 (7.2) | 9 (8.0) | 1 (3.7) | 0.69<sup>d</sup> |
| Rotigotine | 5 (3.6) | 2 (1.8) | 3 (11.1) | 0.050<sup>d</sup> |
| Rasagiline | 29 (20.9) | 24 (21.4) | 5 (18.5) | 0.74<sup>c</sup> |
| Selegiline (oral, sublingual) | 3 (2.1) | 3 (2.7) | 0 (0.00) | 0.99<sup>d</sup> |
| Amantadine (liquid, infusion) | 14 (10.1) | 10 (8.9) | 4 (14.8) | 0.47<sup>d</sup> |
| **Any_medication, n (%)** |  |  |  |  |
|  | 121 (85.8) | 99 (86.8) | 22 (81.5) | 0.54<sup>d</sup> |
| **Current Comorbidities, n (%)** |  |  |  |  |
| ALS | 0 (0.00) | 0 (0.00) | 0 (0.00) |  |
| Alzheimer's disease | 0 (0.00) | 0 (0.00) | 0 (0.00) |  |
| Arrhythmia/Atrial Fibrillation | 9 (6.4) | 8 (7.0) | 1 (3.7) | 0.99<sup>d</sup> |
| Arthritis | 75 (53.2) | 64 (56.1) | 11 (40.7) | 0.15<sup>c</sup> |
| B12 deficiency | 15 (10.6) | 8 (7.0) | 7 (25.9) | **0.010<sup>d</sup>** |
| Cancer | 5 (3.5) | 4 (3.5) | 1 (3.7) | 0.99<sup>d</sup> |
| Congestive Heart Failure | 1 (0.71) | 0 (0.00) | 1 (3.7) | 0.19<sup>d</sup> |
| Crohn's disease/ulcerative colitis | 1 (0.71) | 1 (0.88) | 0 (0.00) | 0.99<sup>d</sup> |
| Depression | 39 (27.7) | 30 (26.3) | 9 (33.3) | 0.46<sup>c</sup> |
| Diabetes Mellitus (adult onset) | 6 (4.3) | 4 (3.5) | 2 (7.4) | 0.32<sup>d</sup> |
| Diabetes Mellitus (childhood onset) | 0 (0.00) | 0 (0.00) | 0 (0.00) |  |
| Gaucher disease | 1 (0.71) | 0 (0.00) | 1 (3.7) | 0.19<sup>d</sup> |
| Gout | 2 (1.4) | 2 (1.8) | 0 (0.00) | 0.99<sup>d</sup> |
| Hearing Loss | 35 (24.8) | 29 (25.4) | 6 (22.2) | 0.73<sup>c</sup> |
| Hip Fracture | 0 (0.00) | 0 (0.00) | 0 (0.00) |  |
| Hyper cholesterolemia (High Cholesterol) | 39 (27.7) | 32 (28.1) | 7 (25.9) | 0.82<sup>c</sup> |
| Hypertension (High Blood Pressure) | 27 (19.1) | 21 (18.4) | 6 (22.2) | 0.65<sup>c</sup> |
| Liver | 2 (1.4) | 2 (1.8) | 0 (0.00) | 0.99<sup>d</sup> |
| Loss of smell | 59 (41.8) | 45 (39.5) | 14 (51.9) | 0.24<sup>c</sup> |
| Lung disease (including emphysema) | 9 (6.4) | 6 (5.3) | 3 (11.1) | 0.37<sup>d</sup> |
| Multiple sclerosis | 0 (0.00) | 0 (0.00) | 0 (0.00) |  |
| Myocardial Infarction (Heart Attack) | 0 (0.00) | 0 (0.00) | 0 (0.00) |  |
| Normal pressure hydrocephalus | 0 (0.00) | 0 (0.00) | 0 (0.00) |  |
| Other | 53 (37.6) | 47 (41.2) | 6 (22.2) | 0.067<sup>c</sup> |
| Peptic ulcer disease | 1 (0.71) | 0 (0.00) | 1 (3.7) | 0.19<sup>d</sup> |
| Peripheral neuropathy | 13 (9.2) | 11 (9.6) | 2 (7.4) | 0.99<sup>d</sup> |
| Peripheral vascular disease | 2 (1.4) | 0 (0.00) | 2 (7.4) | **0.036<sup>d</sup>** |
| PTSD | 8 (5.7) | 5 (4.4) | 3 (11.1) | 0.18<sup>d</sup> |
| Recreational drug use | 8 (5.7) | 7 (6.1) | 1 (3.7) | 0.99<sup>d</sup> |
| Renal insufficiency (kidney disease) | 3 (2.1) | 3 (2.6) | 0 (0.00) | 0.99<sup>d</sup> |
| Seizure, fit, convulsion or unexplained loss of consciousness | 1 (0.71) | 1 (0.88) | 0 (0.00) | 0.99<sup>d</sup> |
| Stroke, mini stroke, CVA (cerebrovascular accident) or TIA (transient ischemic attack) | 1 (0.71) | 0 (0.00) | 1 (3.7) | 0.19<sup>d</sup> |
| Thyroid disease (not cancer, including Grave's disease) | 28 (19.9) | 24 (21.1) | 4 (14.8) | 0.47<sup>c</sup> |
| **UPDRS Score** |  |  |  |  |
| Part 1, mean (sd) | 8.2 (5.7) | 8.0 (5.4) | 9.0 (6.7) | 0.42<sup>a1</sup> |
| Part 2, mean (sd) | 7.1 (6.1) | 6.4 (5.5) | 10.3 (7.2) | **0.002<sup>a1</sup>** |
| Part 3, mean (sd) | 21.4 (12.0) | 17.1 (7.6) | 39.8 (9.0) | **<0.001<sup>a1</sup>** |
| Part 4, mean (sd)* | 3.0 (3.4) | 2.7 (3.2) | 3.9 (4.1) | 0.14<sup>a1</sup> |

\*Data not available for all subjects. Missing values: Genetic Status = 2; Entacapone = 1; Carbidopa, Levodopa, and Entacapone (Stalevo) = 2; Tolcapone (excluding Stalevo) = 2; Apomorphine (injection, infusion, other) = 2; Pramipexole = 2; Ropinirole = 2; Rotigotine = 2; Rasagiline = 2; Selegiline (oral, sublingual) = 1; Amantadine (liquid, infusion) = 2; Part 4 = 19. Statistics presented as Mean (SD), N (column %). p-values: <sup>a1</sup>=t-test, <sup>a2</sup>=Satterthwaite t-test, <sup>c</sup>=Pearson's chi-square test, <sup>d</sup>=Fisher's Exact test.

**Table 1D.** Characteristics of the Study Sample: Stratified by Part IV Severity Groups.

|  | Total | Mild | Moderate/Severe | p-value |
| --- | --- | --- | --- | --- |
| **Sample Size** | 139 | 96 | 43 |  |
| **Age, mean (sd)** | 64.5 (8.9) | 64.3 (9.8) | 65.0 (6.7) | 0.59<sup>a2</sup> |
| **Age at diagnosis, mean (sd)** | 58.0 (10.0) | 59.2 (9.9) | 55.4 (10.0) | **0.039<sup>a1</sup>** |
| **Years since symptom onset, mean (sd)** | 8.9 (6.1) | 7.5 (5.1) | 12.1 (7.1) | **<0.001<sup>a2</sup>** |
| **Years since diagnosis, mean (sd)** | 6.5 (5.3) | 5.1 (3.9) | 9.6 (6.4) | **<0.001<sup>a2</sup>** |
| **Ethnicity, n (%)** |  |  |  | 0.55<sup>d</sup> |
| Hispanic/Latino | 3 (2.2) | 3 (3.1) | 0 (0.00) |  |
| Not Hispanic or Latino | 136 (97.8) | 93 (96.9) | 43 (100.0) |  |
| **Race, n (%)** |  |  |  | 0.90<sup>d</sup> |
| Asian | 1 (0.72) | 1 (1.04) | 0 (0.00) |  |
| Asian, Native Hawaiian or Other Pacific Islander | 1 (0.72) | 1 (1.04) | 0 (0.00) |  |
| Asian, White | 1 (0.72) | 1 (1.04) | 0 (0.00) |  |
| Black or African American | 1 (0.72) | 1 (1.04) | 0 (0.00) |  |
| Black or African American, White | 1 (0.72) | 0 (0.00) | 1 (2.3) |  |
| White | 133 (95.7) | 91 (94.8) | 42 (97.7) |  |
| White, Native American | 1 (0.72) | 1 (1.04) | 0 (0.00) |  |
| **Genetic Status, n (%)\*** |  |  |  | 0.38<sup>d</sup> |
| GBA | 15 (10.9) | 8 (8.4) | 7 (16.3) |  |
| LRRK2 | 10 (7.2) | 5 (5.3) | 5 (11.6) |  |
| PINK1 | 1 (0.72) | 1 (1.05) | 0 (0.00) |  |
| PRKN | 6 (4.3) | 4 (4.2) | 2 (4.7) |  |
| VPS35 | 1 (0.72) | 1 (1.05) | 0 (0.00) |  |
| None | 105 (76.1) | 76 (80.0) | 29 (67.4) |  |
| **Medications, n (%)** |  |  |  |  |
| Levodopa | 115 (82.7) | 78 (81.3) | 37 (86.0) | 0.49<sup>c</sup> |
| Entacapone | 3 (2.2) | 2 (2.1) | 1 (2.3) | 0.99<sup>d</sup> |
| Carbidopa, Levodopa, and Entacapone (Stalevo) | 12 (8.8) | 6 (6.4) | 6 (14.0) | 0.19<sup>d</sup> |
| Tolcapone (excluding Stalevo) | 0 (0.00) | 0 (0.00) | 0 (0.00) |  |
| Apomorphine (injection, infusion, other) | 0 (0.00) | 0 (0.00) | 0 (0.00) |  |
| Pramipexole | 21 (15.3) | 14 (14.9) | 7 (16.3) | 0.83<sup>c</sup> |
| Ropinirole | 11 (8.0) | 6 (6.4) | 5 (11.6) | 0.32<sup>d</sup> |
| Rotigotine | 6 (4.4) | 2 (2.1) | 4 (9.3) | 0.077<sup>d</sup> |
| Rasagiline | 32 (23.4) | 23 (24.5) | 9 (20.9) | 0.65<sup>c</sup> |
| Selegiline (oral, sublingual) | 3 (2.2) | 2 (2.1) | 1 (2.3) | 0.99<sup>d</sup> |
| Amantadine (liquid, infusion) | 16 (11.7) | 8 (8.5) | 8 (18.6) | 0.088<sup>c</sup> |
| **Any_medication, n (%)** |  |  |  |  |
|  | 135 (97.1) | 93 (96.9) | 42 (97.7) | 0.99<sup>d</sup> |
| **Current Comorbidities, n (%)** |  |  |  |  |
| ALS | 0 (0.00) | 0 (0.00) | 0 (0.00) |  |
| Alzheimer's disease | 0 (0.00) | 0 (0.00) | 0 (0.00) |  |
| Arrhythmia/Atrial Fibrillation | 7 (5.0) | 5 (5.2) | 2 (4.7) | 0.99<sup>d</sup> |
| Arthritis | 76 (54.7) | 53 (55.2) | 23 (53.5) | 0.85<sup>c</sup> |
| B12 deficiency | 15 (10.8) | 7 (7.3) | 8 (18.6) | 0.073<sup>d</sup> |
| Cancer | 6 (4.3) | 5 (5.2) | 1 (2.3) | 0.67<sup>d</sup> |
| Congestive Heart Failure | 1 (0.72) | 0 (0.00) | 1 (2.3) | 0.31<sup>d</sup> |
| Crohn's disease/ulcerative colitis | 1 (0.72) | 1 (1.04) | 0 (0.00) | 0.99<sup>d</sup> |
| Depression | 40 (28.8) | 22 (22.9) | 18 (41.9) | **0.023<sup>c</sup>** |
| Diabetes Mellitus (adult onset) | 6 (4.3) | 5 (5.2) | 1 (2.3) | 0.67<sup>d</sup> |
| Diabetes Mellitus (childhood onset) | 0 (0.00) | 0 (0.00) | 0 (0.00) |  |
| Gaucher disease | 1 (0.72) | 0 (0.00) | 1 (2.3) | 0.31<sup>d</sup> |
| Gout | 2 (1.4) | 2 (2.1) | 0 (0.00) | 0.99<sup>d</sup> |
| Hearing Loss | 32 (23.0) | 22 (22.9) | 10 (23.3) | 0.96<sup>c</sup> |
| Hip Fracture | 0 (0.00) | 0 (0.00) | 0 (0.00) |  |
| Hyper cholesterolemia (High Cholesterol) | 40 (28.8) | 29 (30.2) | 11 (25.6) | 0.58<sup>c</sup> |
| Hypertension (High Blood Pressure) | 28 (20.1) | 20 (20.8) | 8 (18.6) | 0.76<sup>c</sup> |
| Liver | 2 (1.4) | 2 (2.1) | 0 (0.00) | 0.99<sup>d</sup> |
| Loss of smell | 62 (44.6) | 37 (38.5) | 25 (58.1) | **0.032<sup>c</sup>** |
| Lung disease (including emphysema) | 11 (7.9) | 6 (6.3) | 5 (11.6) | 0.32<sup>d</sup> |
| Multiple sclerosis | 0 (0.00) | 0 (0.00) | 0 (0.00) |  |
| Myocardial Infarction (Heart Attack) | 0 (0.00) | 0 (0.00) | 0 (0.00) |  |
| Normal pressure hydrocephalus | 0 (0.00) | 0 (0.00) | 0 (0.00) |  |
| Other | 50 (36.0) | 33 (34.4) | 17 (39.5) | 0.56<sup>c</sup> |
| Peptic ulcer disease | 1 (0.72) | 0 (0.00) | 1 (2.3) | 0.31<sup>d</sup> |
| Peripheral neuropathy | 16 (11.5) | 11 (11.5) | 5 (11.6) | 0.99<sup>d</sup> |
| Peripheral vascular disease | 2 (1.4) | 2 (2.1) | 0 (0.00) | 0.99<sup>d</sup> |
| PTSD | 8 (5.8) | 3 (3.1) | 5 (11.6) | 0.11<sup>d</sup> |
| Recreational drug use | 9 (6.5) | 7 (7.3) | 2 (4.7) | 0.72<sup>d</sup> |
| Renal insufficiency (kidney disease) | 4 (2.9) | 2 (2.1) | 2 (4.7) | 0.59<sup>d</sup> |
| Seizure, fit, convulsion or unexplained loss of consciousness | 2 (1.4) | 1 (1.04) | 1 (2.3) | 0.52<sup>d</sup> |
| Stroke, mini stroke, CVA (cerebrovascular accident) or TIA (transient ischemic attack) | 2 (1.4) | 0 (0.00) | 2 (4.7) | 0.094<sup>d</sup> |
| Thyroid disease (not cancer, including Grave's disease) | 30 (21.6) | 21 (21.9) | 9 (20.9) | 0.90<sup>c</sup> |
| **UPDRS Score** |  |  |  |  |
| Part 1, mean (sd) | 8.8 (6.1) | 7.4 (4.7) | 11.9 (7.6) | **<0.001<sup>a2</sup>** |
| Part 2, mean (sd) | 7.8 (6.9) | 6.2 (5.3) | 11.2 (8.7) | **0.001<sup>a2</sup>** |
| Part 3, mean (sd)* | 21.1 (12.1) | 18.8 (9.3) | 26.6 (15.9) | **0.009<sup>a2</sup>** |
| Part 4, mean (sd) | 3.0 (3.4) | 1.1 (1.5) | 7.2 (2.4) | **<0.001<sup>a2</sup>** |

\*Data not available for all subjects. Missing values: Genetic Status = 1; Entacapone = 1; Carbidopa, Levodopa, and Entacapone (Stalevo) = 2; Tolcapone (excluding Stalevo) = 2; Apomorphine (injection, infusion, other) = 2; Pramipexole = 2; Ropinirole = 2; Rotigotine = 2; Rasagiline = 2; Selegiline (oral, sublingual) = 1; Amantadine (liquid, infusion) = 2; Part 3 = 17.
Statistics presented as Mean (SD), N (column %). p-values: <sup>a1</sup>=t-test, <sup>a2</sup>=Satterthwaite t-test, <sup>c</sup>=Pearson's chi-square test, <sup>d</sup>=Fisher's Exact test.

As our previous work stated, the women’s questionnaire is composed of questions that are specific to a woman’s reproductive lifecycle. Participants in this cohort has a mean MoCA score of 27.4 (±2.3) (data not shown), indicating the women who completed this questionnaire had high cognitive aptitude and were able to complete the questionnaire without issues with impairment. Most respondents experienced their onset of PD during menopause (73.7%), compared to those that either experienced PD symptoms during perimenopause (8.8%) or during regular menses (17.5%). Within our cohort, 36.5% had a sex-specific surgery, whether it be a total or partial hysterectomy, oophorectomy, or mastectomy. When asked about contraceptive history, 68% of women in this cohort have used a pill form of birth control, and out of the 304 women, only 22% did not use any form of birth control contraceptive. Interestingly, the majority of respondents have not used hormone replacement therapy (71%). A summary of all responses from the women’s questionnaire can be found in **Table 2**. When asked about pregnancy-related experiences, the majority of respondents have had children (74.3%), although we noticed a higher incidence of infertility experiences (32.2%) or perinatal complications (31.2%) compared to non-PD women. It is important to note that within the United States, the national prevalence rate of history of primary and secondary infertility among women ranges from 15.5 to 19%^33,34^. Lastly, this questionnaire enquires about common comorbidities that respondents might have been diagnosed with during their lifetime. Overall, of the ones listed, hypothyroidism (17.8%) and migraines with no aura (12.8%) were the most common in this cohort.

**Table 2.**
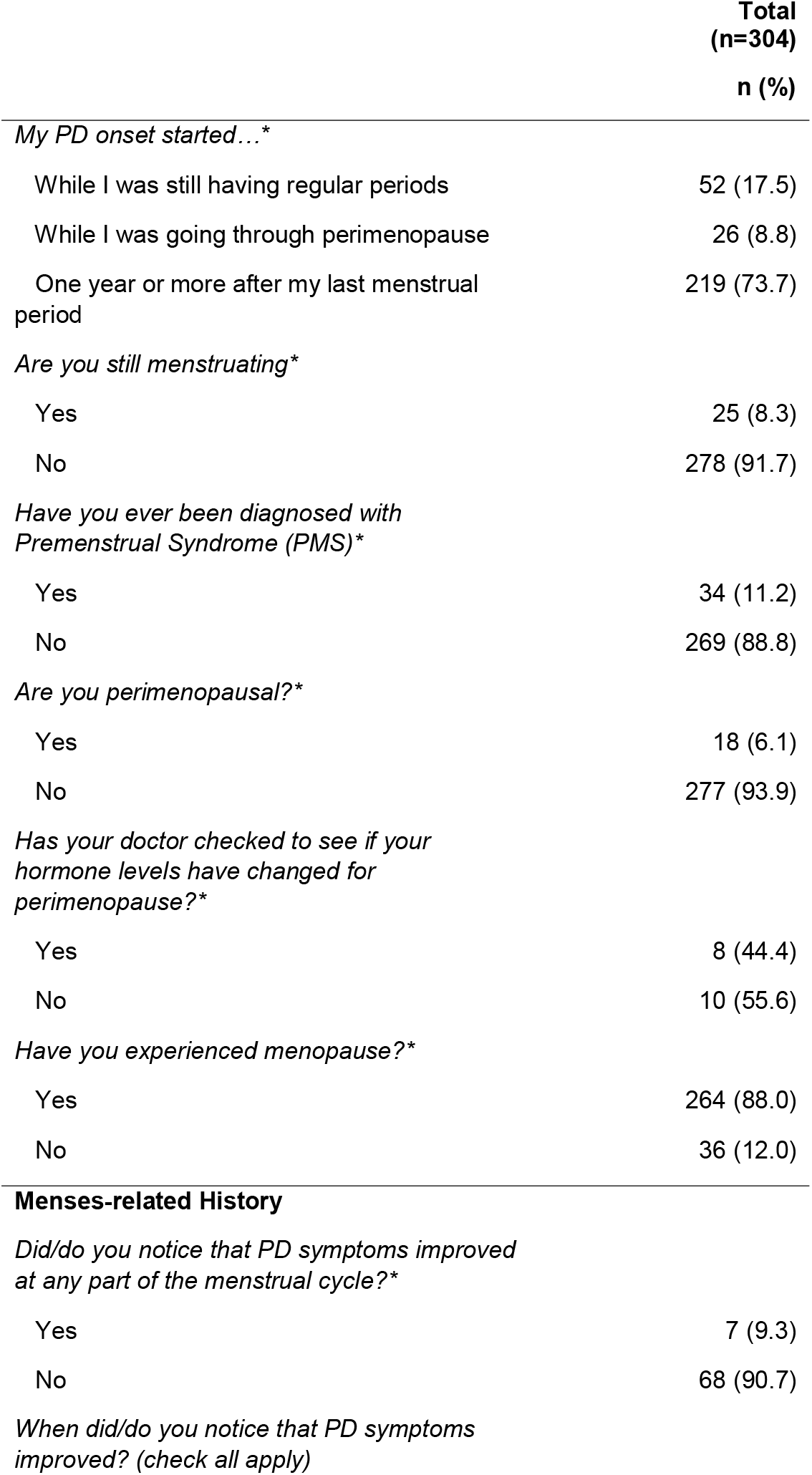
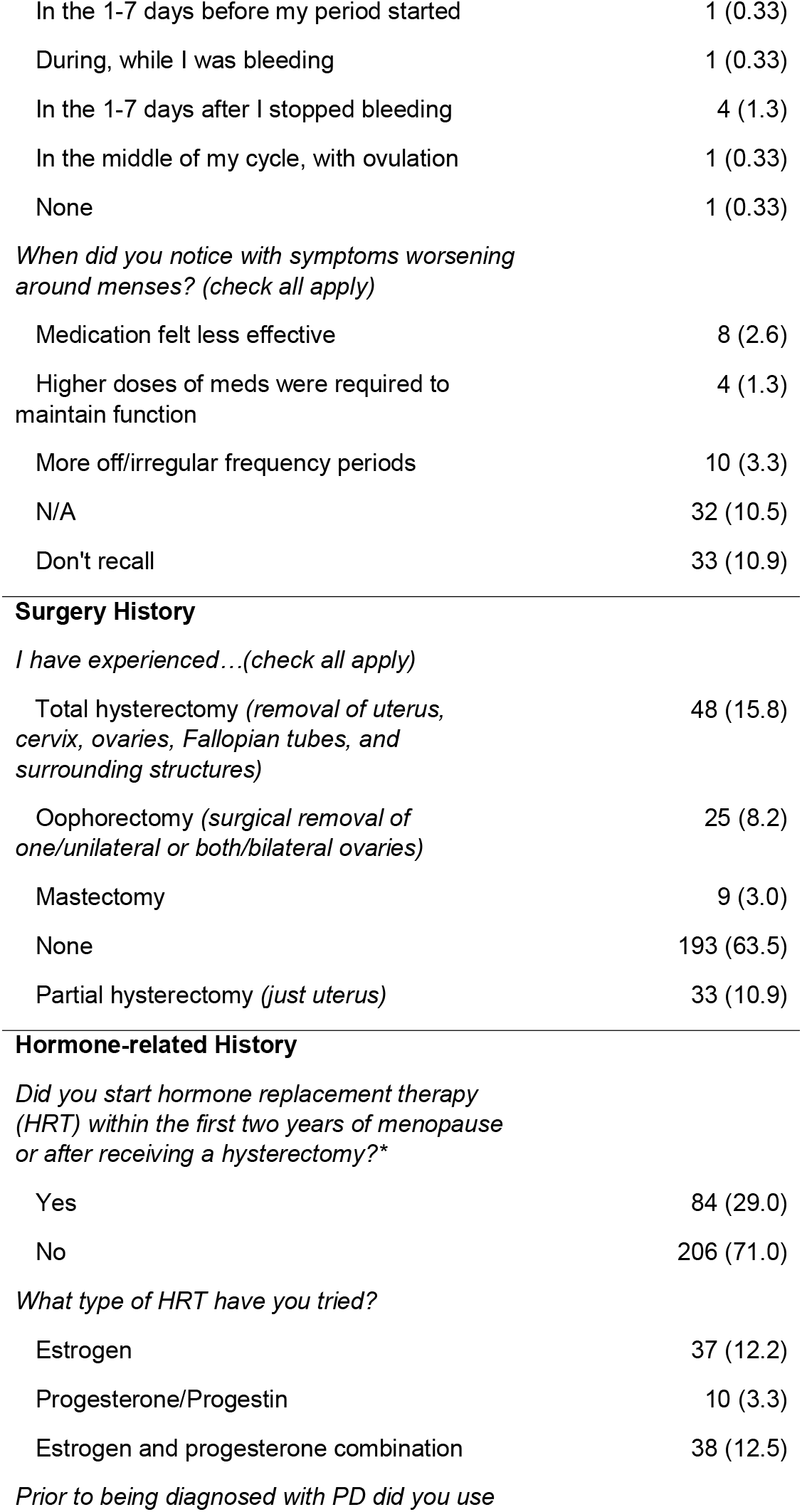
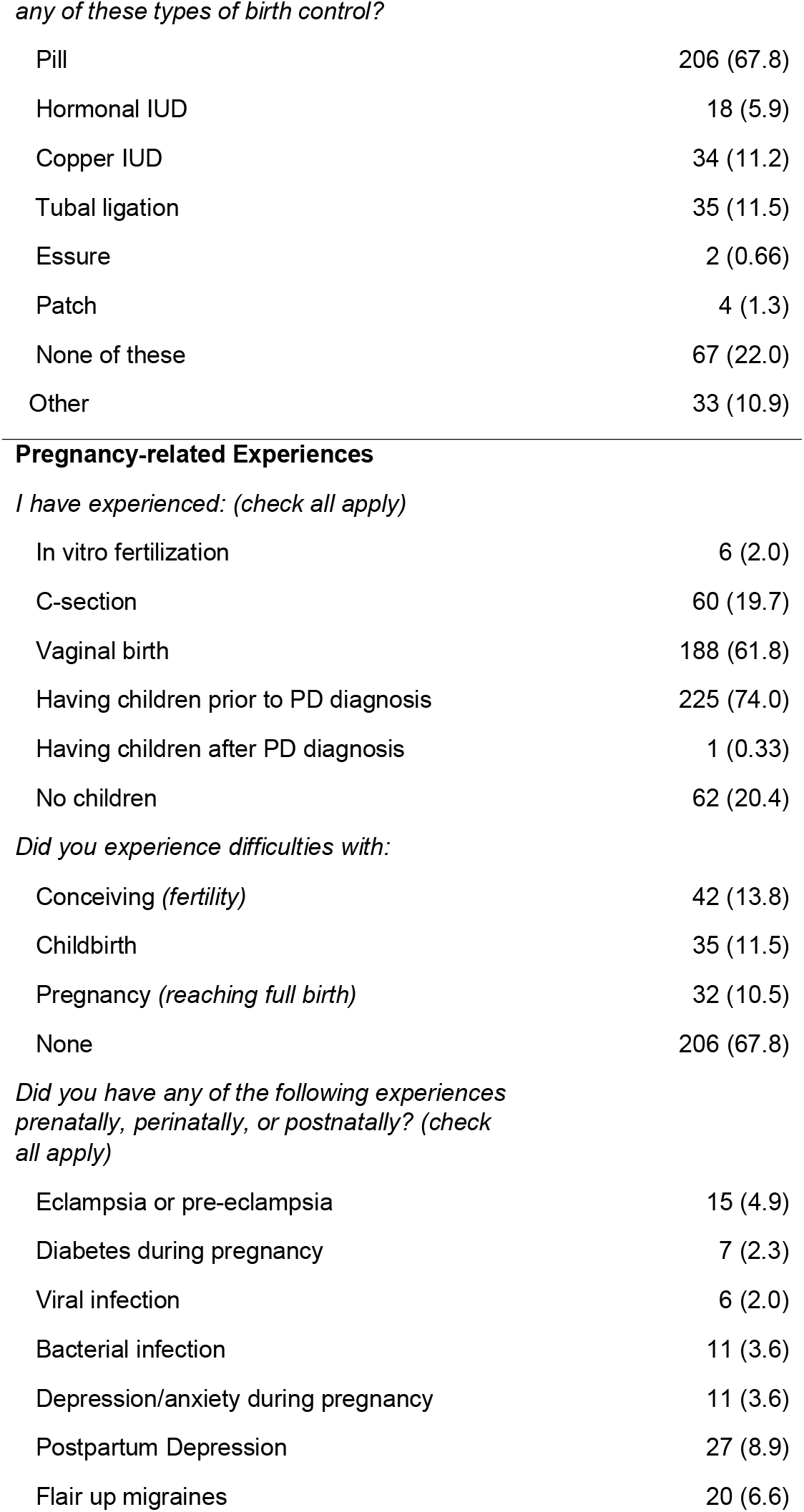
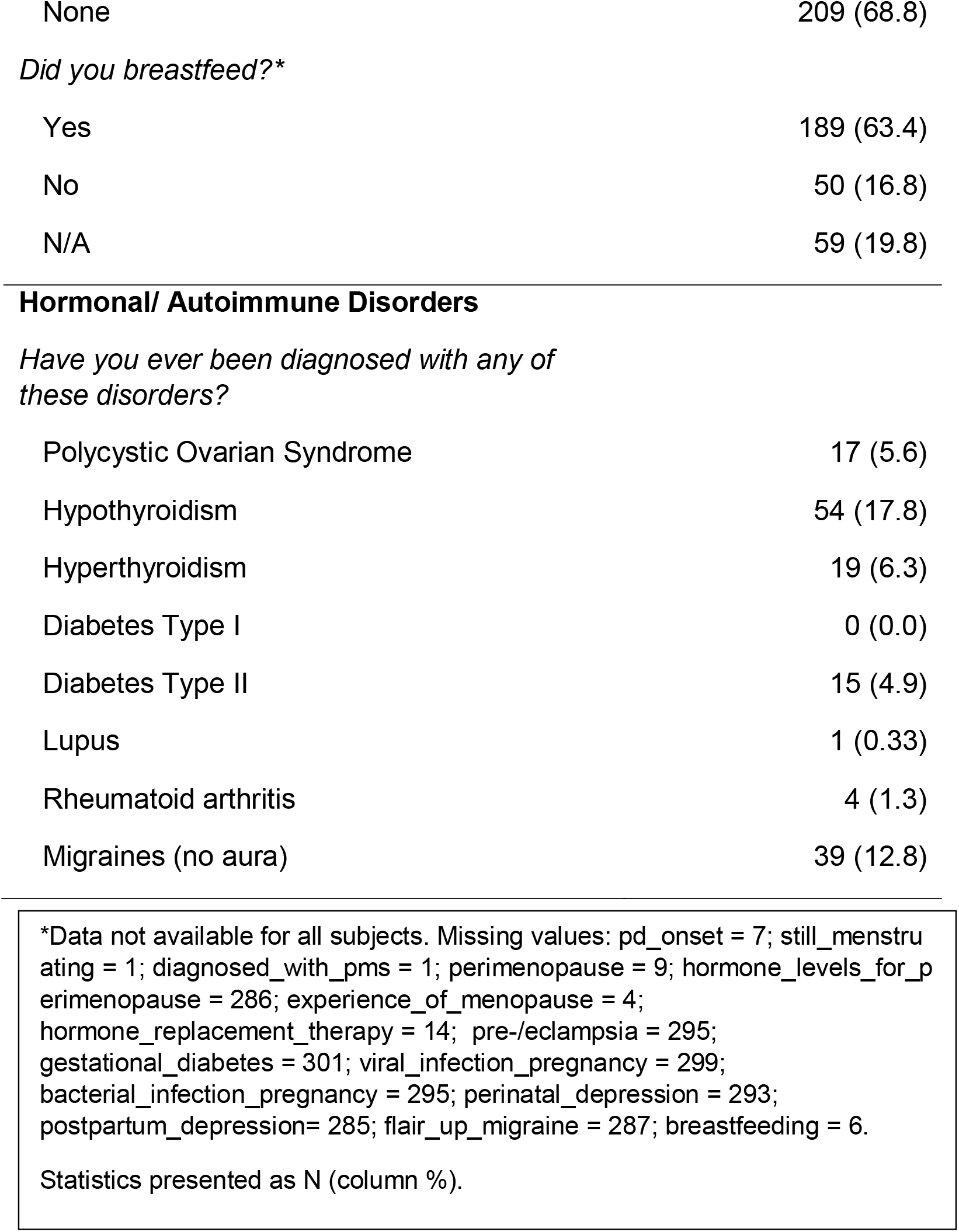
Women’s Health Questionnaire Responses.

|  | **Total**<br>**(n=304)**<br>**n (%)** |
| --- | --- |
| *My PD onset started...\** |  |
| While I was still having regular periods | 52 (17.5) |
| While I was going through perimenopause | 26 (8.8) |
| One year or more after my last menstrual period | 219 (73.7) |
| *Are you still menstruating\** |  |
| Yes | 25 (8.3) |
| No | 278 (91.7) |
| *Have you ever been diagnosed with Premenstrual Syndrome (PMS)\** |  |
| Yes | 34 (11.2) |
| No | 269 (88.8) |
| *Are you perimenopausal?\** |  |
| Yes | 18 (6.1) |
| No | 277 (93.9) |
| *Has your doctor checked to see if your hormone levels have changed for perimenopause?\** |  |
| Yes | 8 (44.4) |
| No | 10 (55.6) |
| *Have you experienced menopause?\** |  |
| Yes | 264 (88.0) |
| No | 36 (12.0) |
| **Menses-related History** |  |
| *Did/do you notice that PD symptoms improved at any part of the menstrual cycle?\** |  |
| Yes | 7 (9.3) |
| No | 68 (90.7) |
| *When did/do you notice that PD symptoms improved? (check all apply)* |  |
| In the 1-7 days before my period started | 1 (0.33) |
| During, while I was bleeding | 1 (0.33) |
| In the 1-7 days after I stopped bleeding | 4 (1.3) |
| In the middle of my cycle, with ovulation | 1 (0.33) |
| None | 1 (0.33) |
| *When did you notice with symptoms worsening around menses? (check all apply)* |  |
| Medication felt less effective | 8 (2.6) |
| Higher doses of meds were required to maintain function | 4 (1.3) |
| More off/irregular frequency periods | 10 (3.3) |
| N/A | 32 (10.5) |
| Don't recall | 33 (10.9) |
| Total hysterectomy (removal of uterus, cervix, ovaries, Fallopian tubes, and surrounding structures) | 48 (15.8) |
| Oophorectomy (surgical removal of one/unilateral or both/bilateral ovaries) | 25 (8.2) |
| Mastectomy | 9 (3.0) |
| None | 193 (63.5) |
| Partial hysterectomy (just uterus) | 33 (10.9) |
| Yes | 84 (29.0) |
| No | 206 (71.0) |
| *What type of HRT have you tried?* |  |
| Estrogen | 37 (12.2) |
| Progesterone/Progestin | 10 (3.3) |
| Estrogen and progesterone combination | 38 (12.5) |
| Pill | 206 (67.8) |
| Hormonal IUD | 18 (5.9) |
| Copper IUD | 34 (11.2) |
| Tubal ligation | 35 (11.5) |
| Essure | 2 (0.66) |
| Patch | 4 (1.3) |
| None of these | 67 (22.0) |
| Other | 33 (10.9) |
| In vitro fertilization | 6 (2.0) |
| C-section | 60 (19.7) |
| Vaginal birth | 188 (61.8) |
| Having children prior to PD diagnosis | 225 (74.0) |
| Having children after PD diagnosis | 1 (0.33) |
| No children | 62 (20.4) |
| *Did you experience difficulties with:* |  |
| Conceiving (*fertility*) | 42 (13.8) |
| Childbirth | 35 (11.5) |
| Pregnancy (*reaching full birth*) | 32 (10.5) |
| None | 206 (67.8) |
| *Did you have any of the following experiences prenatally, perinatally, or postnatally? (check all apply)* |  |
| Eclampsia or pre-eclampsia | 15 (4.9) |
| Diabetes during pregnancy | 7 (2.3) |
| Viral infection | 6 (2.0) |
| Bacterial infection | 11 (3.6) |
| Depression/anxiety during pregnancy | 11 (3.6) |
| Postpartum Depression | 27 (8.9) |
| Flair up migraines | 20 (6.6) |
| None | 209 (68.8) |
| *Did you breastfeed?\** |  |
| Yes | 189 (63.4) |
| No | 50 (16.8) |
| N/A | 59 (19.8) |
| Polycystic Ovarian Syndrome | 17 (5.6) |
| Hypothyroidism | 54 (17.8) |
| Hyperthyroidism | 19 (6.3) |
| Diabetes Type I | 0 (0.0) |
| Diabetes Type II | 15 (4.9) |
| Lupus | 1 (0.33) |
| Rheumatoid arthritis | 4 (1.3) |
| Migraines (no aura) | 39 (12.8) |

\*Data not available for all subjects. Missing values: pd\_onset = 7; still\_menstruating = 1; diagnosed\_with\_pms = 1; perimenopause = 9; hormone\_levels\_for\_perimenopause = 286; experience\_of\_menopause = 4; hormone\_replacement\_therapy = 14; pre-/eclampsia = 295; gestational\_diabetes = 301; viral\_infection\_pregnancy = 299; bacterial\_infection\_pregnancy = 295; perinatal\_depression = 293; postpartum\_depression = 285; flair\_up\_migraine = 287; breastfeeding = 6.
Statistics presented as N (column %).

### Univariate modeling

From our univariate models, we found that certain PD medications, depression, loss of smell, PTSD, B12 deficiency, peripheral neuropathy, and flair-up migraines were associated with a moderate/severe UPDRS score within at least one of the UPDRS Parts. Additionally, for women-specific health factors, we saw that medications feeling less effective during menses, tubal ligation, mastectomy, total hysterectomy, vaginal birth, gestational diabetes, peri and postnatal depression, and perinatal bacterial infection were associated with a moderate/severe PD phenotype. A full summary of the univariate logistic regression with the sample size, OR (95% CI), and p-values can be seen in **Supplemental Table 2**.

### Multivariable modeling

To further assess the role of WSHFs and PD severity, we constructed multivariable logistics regression models using variables that were significant in the univariate models and adjusting for age, disease duration and medication. From Part I, having depression as a current diagnosis was associated with a moderate/severe PD severity phenotype (OR = 9.22 (3.31, 25.71), p<0.001). In terms of pregnancy-related experiences, we found that having a natural (vaginal) birth was significantly associated with the moderate/severe cohort (OR = 4.48 (1.10, 18.20), p = 0.036), and postpartum depression demonstrated an association that did not reach conventional statistical significance (OR = 4.17 (0.91, 19.20), p=0.067). Through our modeling, we noticed that “other comorbidities” was significant as well (OR = 3.91 (1.51, 10,14), p = 0.005). This was gathered as part of PD GENEration’s clinical history, where participants can include specific disorders they were diagnosed with; these comorbidities are listed in **Supplemental File 1**. For Part II, we saw that years since diagnosis was significant (OR = 1.17 (1.09, 1.25), p<0.001), which is expected as Part II assesses motor experiences in daily living. Depression and perinatal depression were significantly associated with the moderate/severe PD phenotype (OR = 2.33 (1.13, 4.80), p=0.022, OR = 6.72 (1.35, 33.43), p=0.020, respectively). In Part III, the motor examination, having a *LRRK2* genotype (OR = 6.74 (1.62, 28.00), p=0.009), total hysterectomy (OR = 5.46 (1.74, 17.18), p=0.004), and being diagnosed with B12 deficiency (OR = 6.22 (1.69, 22.95), p=0.006) were significantly associated with higher PD severity. For Part IV, only years since diagnosis was significant (OR = 1.17 (1.06, 1.29), p=0.002). Overall, the concordance statistic (c-statistic) for these four regression models demonstrated high accuracy for discerning PD severity for women within this cohort (Part I: 0.874, Part II: 0.805, Part III 0.791, and Part IV: 0.782). The univariate variables in the multivariable regressions with the c-statistic, OR (95% CI), and p-values are summarized in **Table 3**.

**Table 3A.**
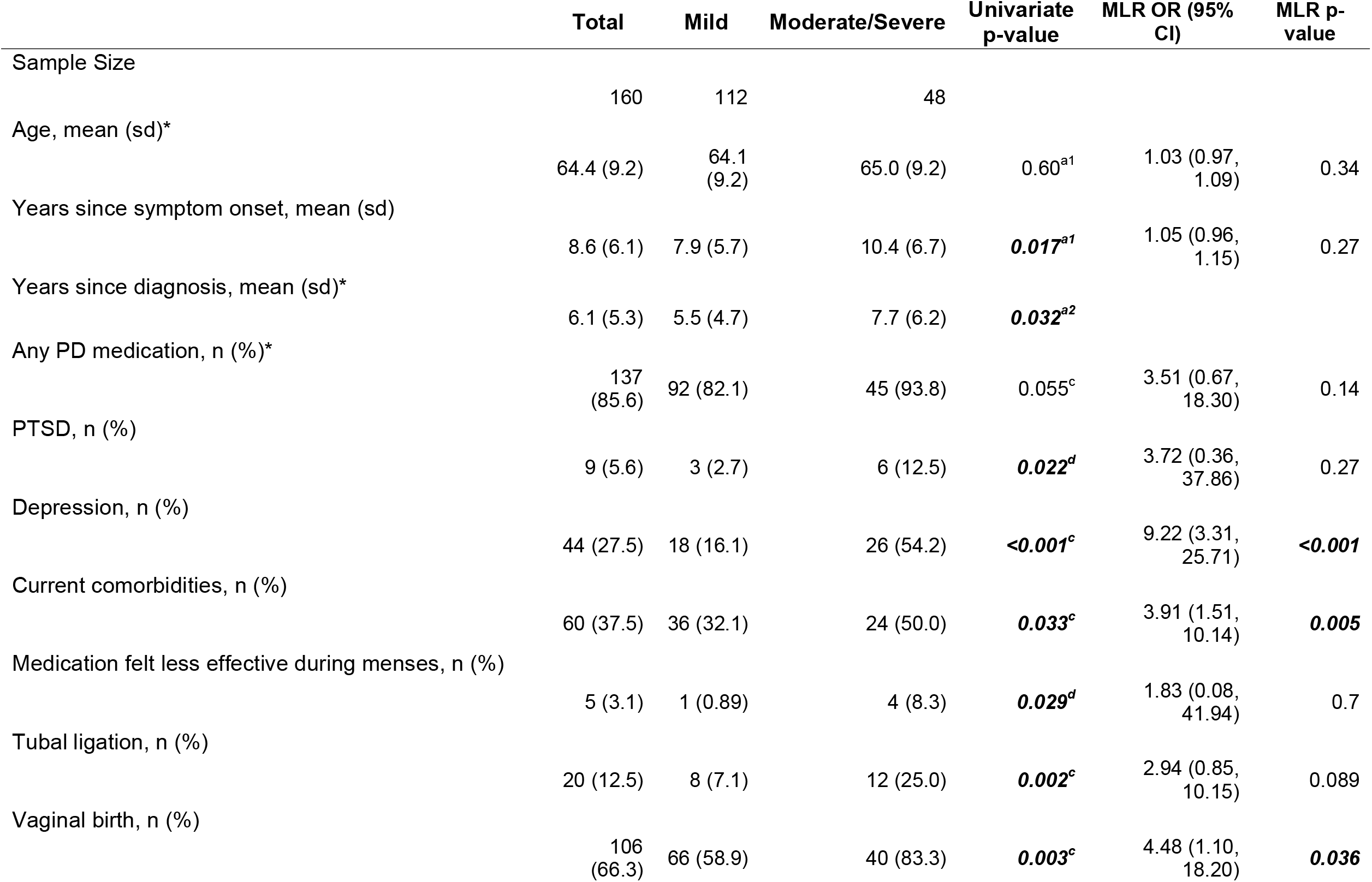
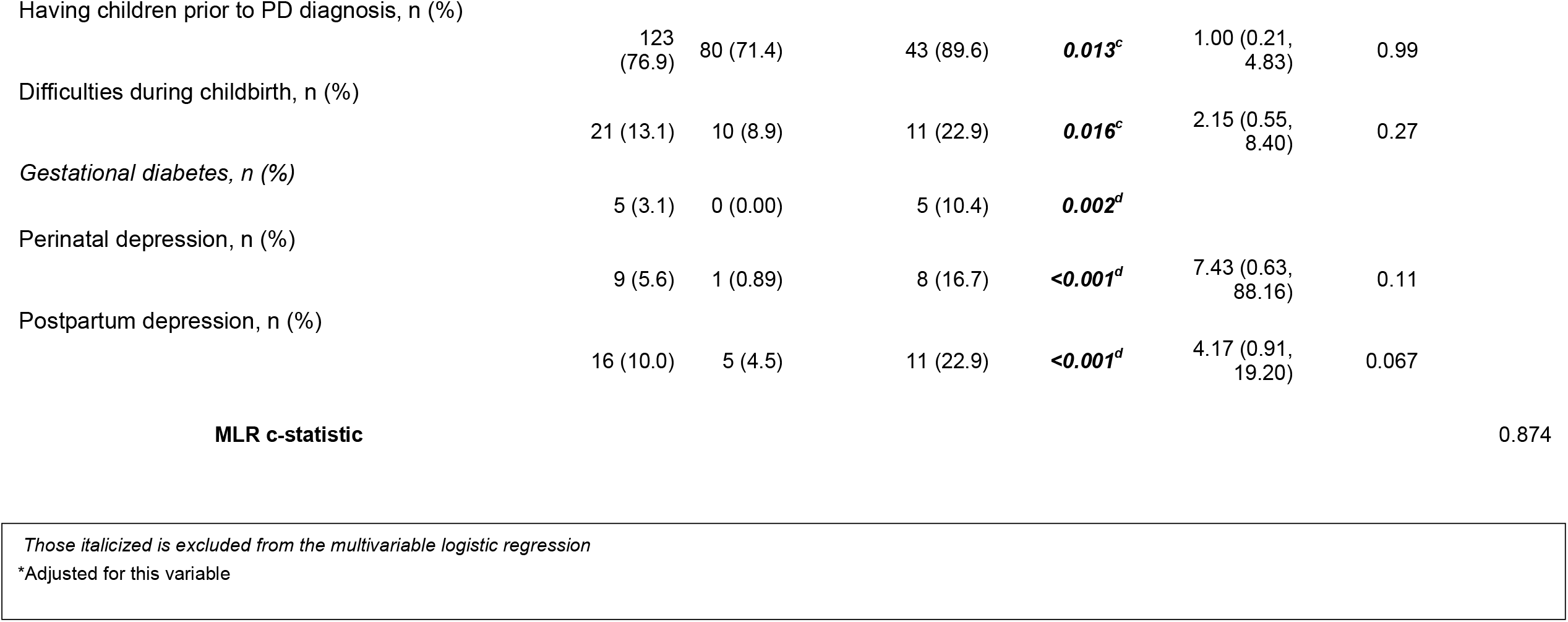
Summary of Part I Multivariable Logistic Regression (MLR) Results

|  | Total | Mild | Moderate/Severe | Univariate<br>p-value | MLR OR (95%<br>CI) | MLR p-value |
| --- | --- | --- | --- | --- | --- | --- |
| Sample Size | 160 | 112 | 48 |  |  |  |
| Age, mean (sd)* | 64.4 (9.2) | 64.1<br>(9.2) | 65.0 (9.2) | 0.60<sup>a1</sup> | 1.03 (0.97,<br>1.09) | 0.34 |
| Years since symptom onset, mean (sd) | 8.6 (6.1) | 7.9 (5.7) | 10.4 (6.7) | **0.017<sup>a1</sup>** | 1.05 (0.96,<br>1.15) | 0.27 |
| Years since diagnosis, mean (sd)* | 6.1 (5.3) | 5.5 (4.7) | 7.7 (6.2) | **0.032<sup>a2</sup>** |  |  |
| Any PD medication, n (%)* | 137<br>(85.6) | 92 (82.1) | 45 (93.8) | 0.055<sup>c</sup> | 3.51 (0.67,<br>18.30) | 0.14 |
| PTSD, n (%) | 9 (5.6) | 3 (2.7) | 6 (12.5) | **0.022<sup>d</sup>** | 3.72 (0.36,<br>37.86) | 0.27 |
| Depression, n (%) | 44 (27.5) | 18 (16.1) | 26 (54.2) | **<0.001<sup>c</sup>** | 9.22 (3.31,<br>25.71) | **<0.001** |
| Current comorbidities, n (%) | 60 (37.5) | 36 (32.1) | 24 (50.0) | **0.033<sup>c</sup>** | 3.91 (1.51,<br>10.14) | **0.005** |
| Medication felt less effective during menses, n (%) | 5 (3.1) | 1 (0.89) | 4 (8.3) | **0.029<sup>d</sup>** | 1.83 (0.08,<br>41.94) | 0.7 |
| Tubal ligation, n (%) | 20 (12.5) | 8 (7.1) | 12 (25.0) | **0.002<sup>c</sup>** | 2.94 (0.85,<br>10.15) | 0.089 |
| Vaginal birth, n (%) | 106<br>(66.3) | 66 (58.9) | 40 (83.3) | **0.003<sup>c</sup>** | 4.48 (1.10,<br>18.20) | **0.036** |

Having children prior to PD diagnosis, n (%)
|  |  |  |  |  |  |
| --- | --- | --- | --- | --- | --- |
| 123<br>(76.9) | 80 (71.4) | 43 (89.6) | ***0.013<sup>c</sup>*** | 1.00 (0.21,<br>4.83) | 0.99 |

Difficulties during childbirth, n (%)
|  |  |  |  |  |  |
| --- | --- | --- | --- | --- | --- |
| 21 (13.1) | 10 (8.9) | 11 (22.9) | ***0.016<sup>c</sup>*** | 2.15 (0.55,<br>8.40) | 0.27 |

*Gestational diabetes, n (%)*
|  |  |  |  |
| --- | --- | --- | --- |
| 5 (3.1) | 0 (0.00) | 5 (10.4) | ***0.002<sup>d</sup>*** |

Perinatal depression, n (%)
|  |  |  |  |  |  |
| --- | --- | --- | --- | --- | --- |
| 9 (5.6) | 1 (0.89) | 8 (16.7) | ***<0.001<sup>d</sup>*** | 7.43 (0.63,<br>88.16) | 0.11 |

Postpartum depression, n (%)
|  |  |  |  |  |  |
| --- | --- | --- | --- | --- | --- |
| 16 (10.0) | 5 (4.5) | 11 (22.9) | ***<0.001<sup>d</sup>*** | 4.17 (0.91,<br>19.20) | 0.067 |

**Table 3B.**
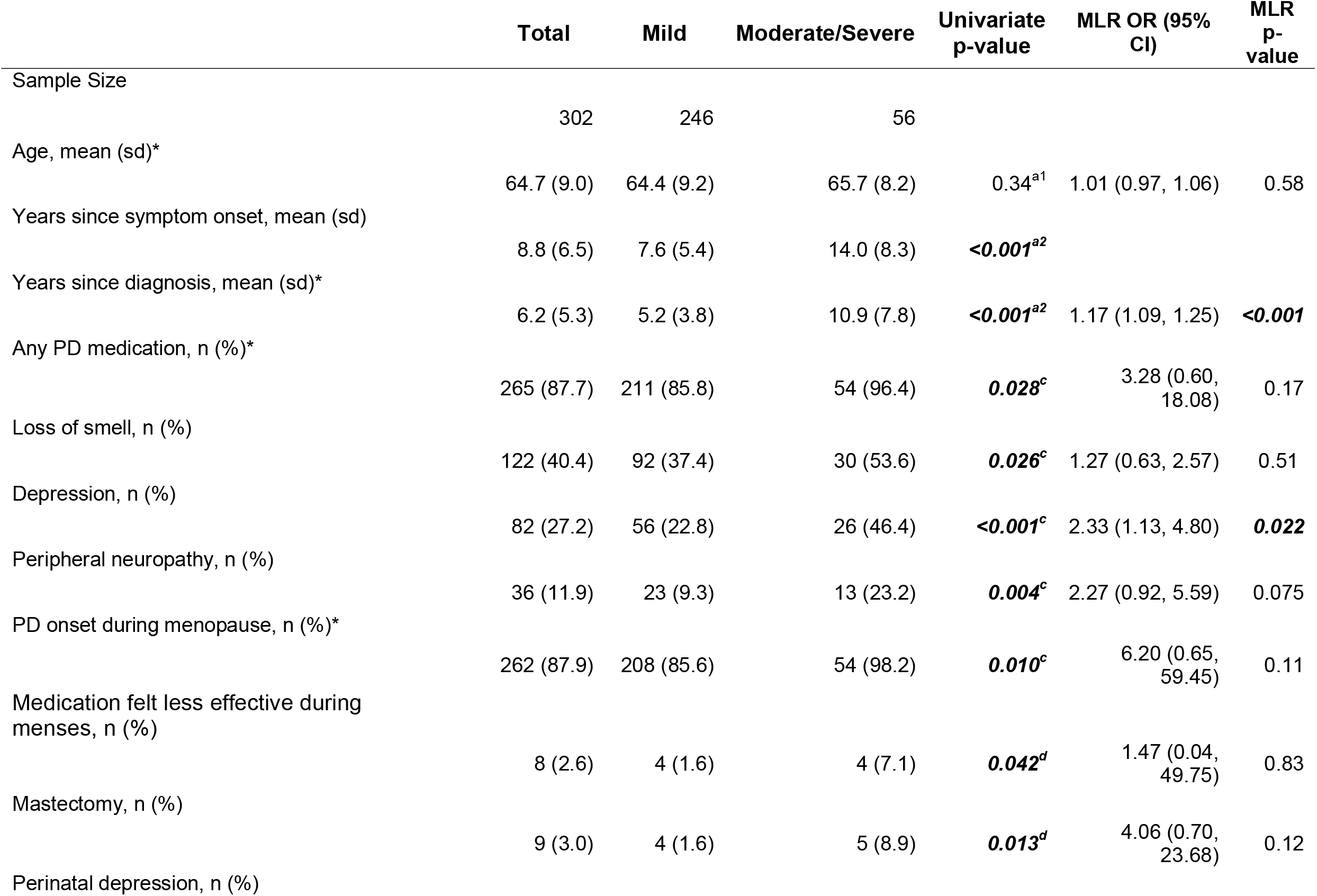
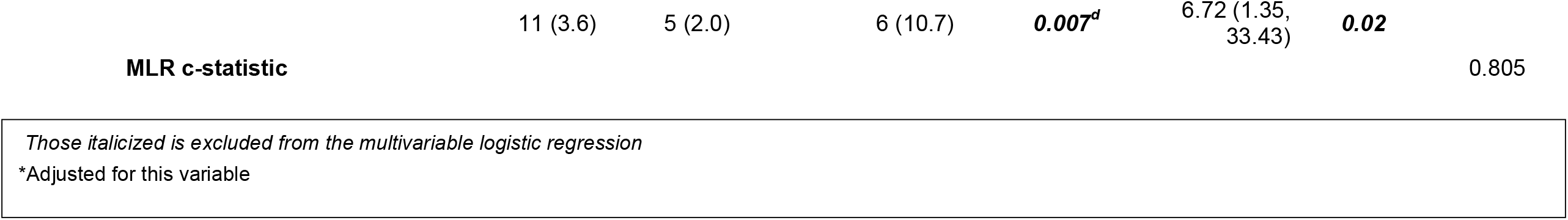
Summary of Part II Multivariable Logistic Regression (MLR) Results

|  | Total | Mild | Moderate/Severe | Univariate<br>p-value | MLR OR (95%<br>CI) | MLR<br>p-value |
| --- | --- | --- | --- | --- | --- | --- |
| Sample Size | 302 | 246 | 56 |  |  |  |
| Age, mean (sd)* | 64.7 (9.0) | 64.4 (9.2) | 65.7 (8.2) | 0.34<sup>a1</sup> | 1.01 (0.97, 1.06) | 0.58 |
| Years since symptom onset, mean (sd) | 8.8 (6.5) | 7.6 (5.4) | 14.0 (8.3) | **<0.001<sup>a2</sup>** |  |  |
| Years since diagnosis, mean (sd)* | 6.2 (5.3) | 5.2 (3.8) | 10.9 (7.8) | **<0.001<sup>a2</sup>** | 1.17 (1.09, 1.25) | **<0.001** |
| Any PD medication, n (%)* | 265 (87.7) | 211 (85.8) | 54 (96.4) | **0.028<sup>c</sup>** | 3.28 (0.60,<br>18.08) | 0.17 |
| Loss of smell, n (%) | 122 (40.4) | 92 (37.4) | 30 (53.6) | **0.026<sup>c</sup>** | 1.27 (0.63, 2.57) | 0.51 |
| Depression, n (%) | 82 (27.2) | 56 (22.8) | 26 (46.4) | **<0.001<sup>c</sup>** | 2.33 (1.13, 4.80) | **0.022** |
| Peripheral neuropathy, n (%) | 36 (11.9) | 23 (9.3) | 13 (23.2) | **0.004<sup>c</sup>** | 2.27 (0.92, 5.59) | 0.075 |
| PD onset during menopause, n (%)* | 262 (87.9) | 208 (85.6) | 54 (98.2) | **0.010<sup>c</sup>** | 6.20 (0.65,<br>59.45) | 0.11 |
| Medication felt less effective during<br>menses, n (%) | 8 (2.6) | 4 (1.6) | 4 (7.1) | **0.042<sup>d</sup>** | 1.47 (0.04,<br>49.75) | 0.83 |
| Mastectomy, n (%) | 9 (3.0) | 4 (1.6) | 5 (8.9) | **0.013<sup>d</sup>** | 4.06 (0.70,<br>23.68) | 0.12 |
| Perinatal depression, n (%) |  |  |  |  |  |  |

|  |  |  |  |  |  |  |  |
| --- | --- | --- | --- | --- | --- | --- | --- |
|  | 11 (3.6) | 5 (2.0) | 6 (10.7) | ***0.007<sup>d</sup>*** | 6.72 (1.35,<br>33.43) | ***0.02*** |  |
| **MLR c-statistic** |  |  |  |  |  |  | 0.805 |

*Those italicized is excluded from the multivariable logistic regression* \*Adjusted for this variable

**Table 3C.** Summary of Part III Multivariable Logistic Regression (MLR) Results

|  | Total | Mild | Moderate/Severe | Univariate<br>p-value | MLR OR (95%<br>CI) | MLR p-value |
| --- | --- | --- | --- | --- | --- | --- |
| Sample Size | 141 | 114 | 27 |  |  |  |
| Age, mean (sd)* | 64.1 (8.7) | 64.0 (8.8) | 64.6 (8.5) | 0.72<sup>a1</sup> | 1.00 (0.94, 1.05) | 0.87 |
| Years since symptom onset, mean (sd) | 8.8 (6.0) | 8.5 (5.6) | 10.1 (7.2) | 0.19<sup>a1</sup> |  |  |
| Years since diagnosis, mean (sd)* | 6.2 (5.2) | 5.9 (4.6) | 7.8 (7.2) | 0.19<sup>a2</sup> | 1.07 (0.99, 1.17) | 0.1 |
| Any PD medication, n (%) | 121 (85.8) | 99 (86.8) | 22 (81.5) | 0.54<sup>d</sup> | 0.44 (0.12, 1.58) | 0.21 |
| LRRK2, n (%) | 11 (7.8) | 6 (5.3) | 5 (18.5) | **0.036<sup>d</sup>** | 6.74 (1.62, 28.00) | **0.009** |
| B12 deficiency, n (%) | 15 (10.6) | 8 (7.0) | 7 (25.9) | **0.010<sup>d</sup>** | 6.22 (1.69, 22.95) | **0.006** |
| Total hysterectomy, n (%) | 24 (17.0) | 15 (13.2) | 9 (33.3) | **0.021<sup>d</sup>** | 5.46 (1.74, 17.18) | **0.004** |
| **MLR c-statistic** |  |  |  |  |  | 0.791 |

*Those italicized is excluded from the multivariable logistic regression*
\*Adjusted for this variable

**Table 3D.**
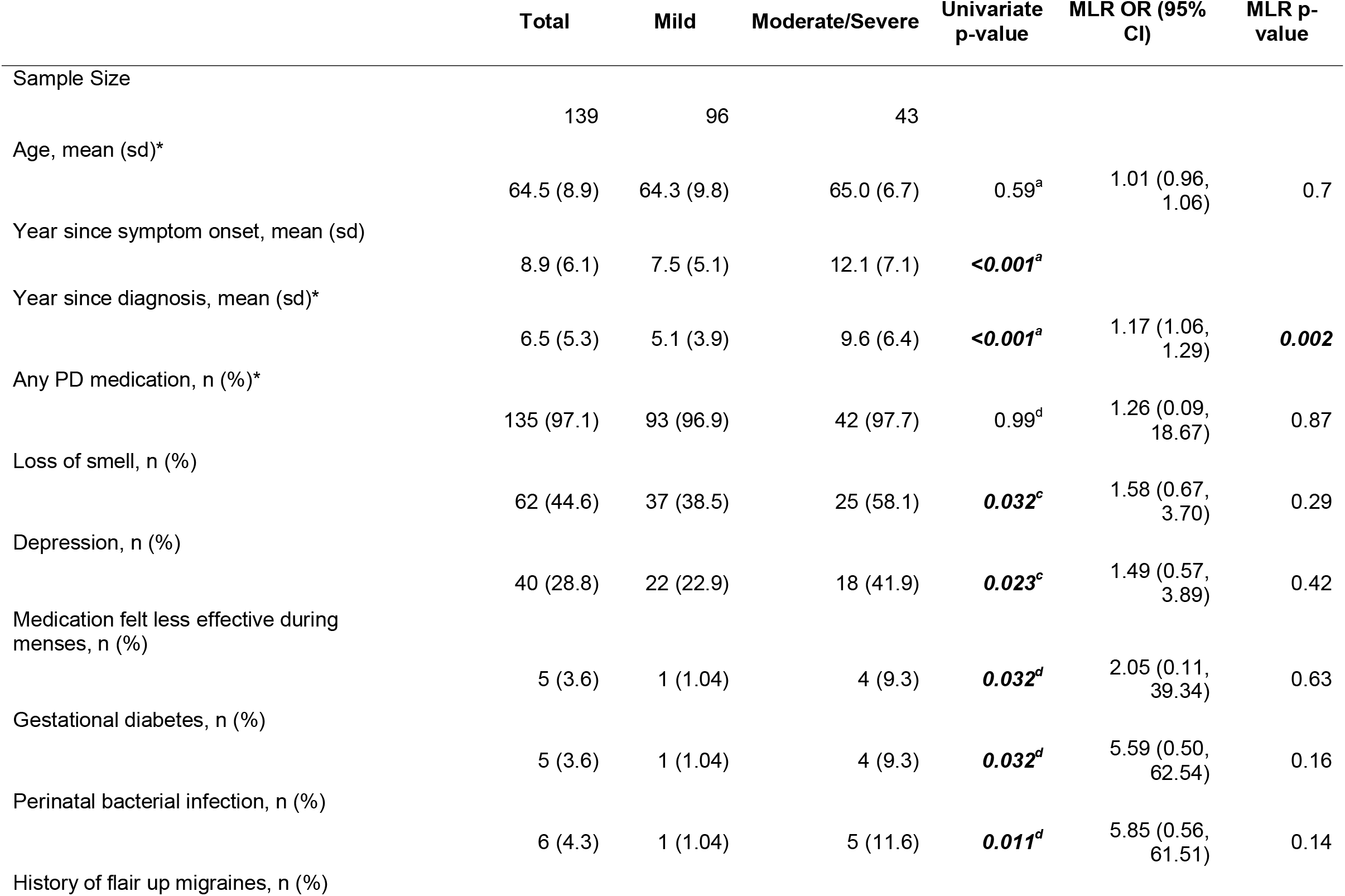
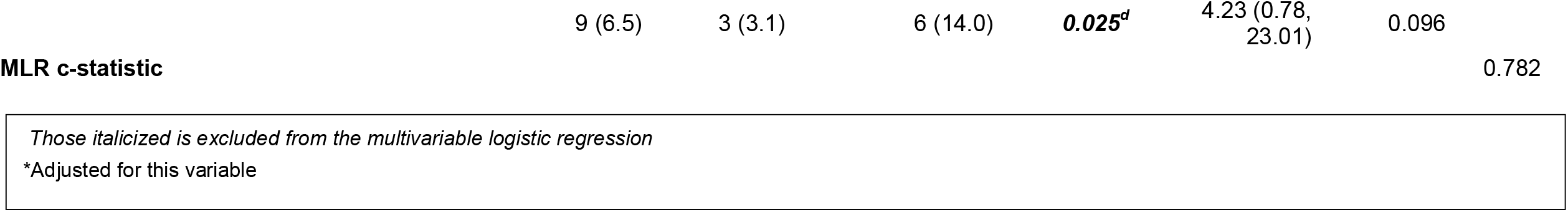
Summary of Part IV Multivariable Logistic Regression (MLR) Results

|  | Total | Mild | Moderate/Severe | Univariate<br>p-value | MLR OR (95%<br>CI) | MLR p-value |
| --- | --- | --- | --- | --- | --- | --- |
| Sample Size | 139 | 96 | 43 |  |  |  |
| Age, mean (sd)* | 64.5 (8.9) | 64.3 (9.8) | 65.0 (6.7) | 0.59<sup>a</sup> | 1.01 (0.96,<br>1.06) | 0.7 |
| Year since symptom onset, mean (sd) | 8.9 (6.1) | 7.5 (5.1) | 12.1 (7.1) | **<0.001<sup>a</sup>** |  |  |
| Year since diagnosis, mean (sd)* | 6.5 (5.3) | 5.1 (3.9) | 9.6 (6.4) | **<0.001<sup>a</sup>** | 1.17 (1.06,<br>1.29) | **0.002** |
| Any PD medication, n (%)* | 135 (97.1) | 93 (96.9) | 42 (97.7) | 0.99<sup>d</sup> | 1.26 (0.09,<br>18.67) | 0.87 |
| Loss of smell, n (%) | 62 (44.6) | 37 (38.5) | 25 (58.1) | **0.032<sup>c</sup>** | 1.58 (0.67,<br>3.70) | 0.29 |
| Depression, n (%) | 40 (28.8) | 22 (22.9) | 18 (41.9) | **0.023<sup>c</sup>** | 1.49 (0.57,<br>3.89) | 0.42 |
| Medication felt less effective during<br>menses, n (%) | 5 (3.6) | 1 (1.04) | 4 (9.3) | **0.032<sup>d</sup>** | 2.05 (0.11,<br>39.34) | 0.63 |
| Gestational diabetes, n (%) | 5 (3.6) | 1 (1.04) | 4 (9.3) | **0.032<sup>d</sup>** | 5.59 (0.50,<br>62.54) | 0.16 |
| Perinatal bacterial infection, n (%) | 6 (4.3) | 1 (1.04) | 5 (11.6) | **0.011<sup>d</sup>** | 5.85 (0.56,<br>61.51) | 0.14 |
| History of flair up migraines, n (%) |  |  |  |  |  |  |

|  |  |  |  |  |  |  |  |
| --- | --- | --- | --- | --- | --- | --- | --- |
|  | 9 (6.5) | 3 (3.1) | 6 (14.0) | ***0.025<sup>d</sup>*** | 4.23 (0.78,<br>23.01) | 0.096 |  |
| **MLR c-statistic** |  |  |  |  |  |  | 0.782 |

*Those italicized is excluded from the multivariable logistic regression*
\*Adjusted for this variable

## Discussion

This study is the first of its nature to look at WSHFs concerning PD severity in women within the US through the deployment of a women-specific health questionnaire. We leveraged PD GENEration’s unique dataset of clinical and genetic health data from PD participants, coupled with the questionnaire responses, to develop multivariable logistic models. Using Goetz et al.’s thresholds, we created mild versus moderate/severe groups for PD phenotype for each subpart of the UPDRS assessment. Our analysis found that WSHFs, such as perinatal depression, delivery of children through vaginal birth, and history of a total hysterectomy, were associated with a severe PD phenotype. Other factors, including major depression disorder, *LRRK2* mutation carrier status, and B12 deficiency, were seen amongst women with a more severe phenotype compared to those with a mild PD phenotype. This study sets the stage for acknowledging the role WSHFs have in the severity of PD. While there have been studies looking at PD risk and some WSHFs, they overlook the growing population of women who have already been diagnosed with PD and thus have a declining health-related quality of life^7,17–19,35–43^.

Through our multivariable logistic regression models, we found that certain variables were associated with a moderate/severe PD phenotype within each subpart of the UPDRS assessment. There has been a great emphasis on the associations between depression and PD. Depression (major depressive disorder) appeared in Parts I and II. While our study cannot definitively say if it was an early-onset symptom of PD or if it was due to a more severe PD phenotype, depression appeared as a comorbidity of PD. Literature has shown that depression has been a prodromal symptom seen more commonly in women than in men, and as a result, women are more often misdiagnosed compared to men^16,44,45^. Moreover, perinatal depression was associated with a moderate/severe UPDRS Part II phenotype. Due to PD being more prevalent among older women, pregnancy-related factors are often neglected in PD studies, despite pregnancy having short-term and long-term effects on maternal health^46,47^. The few studies that have looked at pregnancy and PD, they have focused on women with young-onset PD, who have a different profile than typical women with PD^48,49^. Thus, further studies will be needed to determine if perinatal factors like depression influence PD etiology in women.

Pregnancy has been shown to cause neurological changes in women for them to transition to a more maternal mindset^50–53^. Thus, one might hypothesize that complications that can arise during this phase, such as perinatal depression, could alter the physiological adaptions required for the transition to motherhood, resulting in a woman being predisposed to having PD and, even more so, a more severe PD phenotype. While this study has found major depressive disorder and perinatal depression to be associated with a more severe PD phenotype, more studies are needed to parse out the exact mechanisms of these disorders in PD etiology. Furthermore, within this cohort, natural delivery of a child was significantly associated with a moderate/severe phenotype. In 2020, about 69% of American women experienced a natural delivery, but the long-term implications of natural and cesarean deliveries remain unclear for maternal health and aging^54,55^. Additionally, due to a low percentage of women having children while diagnosed with PD, little is known about if cesarean or vaginal births influence the progression and severity of PD. Although one study found little research regarding PD and pregnancy, after surveying clinicians, it observed that being diagnosed with PD would not impact a woman’s ability to have a vaginal delivery and cesarean sections should be reserved as an alternative method of delivery^56^. Other studies align with this notion and found little evidence for women with PD to prefer cesarean over vaginal delivery, regardless of if a woman was currently on PD treatment and medication or not^57,58^. Thus, while our study demonstrates that having a vaginal delivery was associated with a more severe PD phenotype, this is not evident in other studies, and further studies must consider if the method of child delivery impacts PD etiology.

This study found that carrying a *LRRK2* mutation was associated with a moderate/severe phenotype in Part III of UPDRS. *LRRK2* (leucine-rich repeat kinase 2) gene encodes for a ROCO family protein and is a major contributor to PD risk, as mutations in *LRRK2* have been linked to dopaminergic nerve cell death and impaired dopamine neurotransmission^59^. *LRRK2* mutations can cause autosomal dominant PD, which has been present in up to 40% of familial PD cases in certain ethnic groups^60^. Differences in PD risk and presentation between men and women with *LRRK2* G2019S mutations have been well characterized in the field^61–63^. Overall, women have a relatively higher incidence of having a *LRRK2* G2019S mutation compared to men, and studies observed differences in presentation and medication dosages between men and women with *LRRK2* mutation status^61,64^. Our study showed that the *LRRK2* mutation was associated with a more severe PD phenotype, but this should be considered with caution as our sample size was small (n=11). Further studies are needed to determine if genetic mutations in women cause a more severe motor severity, as only one study has briefly mentioned no differences in severity between women with *LRRK2* carrier status and those without^61^. Since many studies around *LRRK2* focus on the p.G2019S mutation, our cohort had a small representation of this genetic variant n = 10; thus, our findings suggest that the severity of PD for women carrying *LRRK2* mutation could stem from variants that are not p.G2019S such as p.A1215T, p.R1628P, and p.M167R variants.

B12 deficiency was another variable that was significantly associated with a higher UPDRS Part III subscore. Overall, B12 has been characterized as an important vitamin in the maintenance of PD nutrition. Studies have shown that low B12 is linked to cognitive impairment, higher Hoehn and Yahr scores, and dementia-related symptoms^65,66^. B12 deficiency is commonly seen in pregnant women, vegetarians, the elderly, and those who have neurodegenerative disorders^67,68^. The DATATOP study, a large two-year study of patients with early-onset PD, demonstrated that a more rapid progression of PD symptoms occurred in those with lower B12 levels than those with higher levels. This study was foundational in finding that monitoring B12 levels may delay the onset of PD severity^69,70^. Therefore, it is reasonable to observe that those with a moderate/severe phenotype in our study had a B12 deficiency compared to those with mild PD severity. While this association appeared in Part III (motor examination), interestingly, there was also a trend of significance in Part II (motor experiences of daily living) of this subcohort (OR = 2.27 (1.13,4.80) p=0.075). Since literature has suggested that B12 deficiency has been associated with peripheral neuropathy^71,72^, this observation of statistical significance in our cohort suggests a possible connection between B12 deficiency and the trend of peripheral neuropathy in women with a more severe PD phenotype.

In our study, we defined total hysterectomies as the removal of the entire uterus, cervix, and ovaries/fallopian tubes. Although the removal of the ovaries and fallopian tubes is not included in the definition of total hysterectomies, they are often included in the procedure based on the patient’s medical history. Total hysterectomies were significantly associated with a more severe PD phenotype in the PD GENEration questionnaire respondents. Hysterectomies are one of the most commonly performed procedures done in women (second after cesarean sections), and they treat gynecologic malignancies and benign gynecologic diseases^73,74^. About 1 in 3 women in the United States have a hysterectomy by the age of 60^75^, and one study approximates that about 600,000 of these procedures are performed annually in the United States^76^. Removal of the ovaries leads to patients taking exogenous hormones, which have been studied in relation to PD risk. However, across these studies, there is no conclusive answer to if exogenous hormones contribute to PD risk. One study found that hysterectomies, with or without unilateral oophorectomy (removal of one ovary), were significantly associated with PD risk^35^. Other studies found similar results with women being at a higher risk for PD if they either had unilateral or bilateral oophorectomies before experiencing menopause or if they used estrogen alone after a hysterectomy^18,19,40^. On the other hand, one study found that anemia, a condition that is more common in women and can lead to hysterectomies, was associated with PD risk in women, even after adjusting for hysterectomies, which indicates that the procedure is not a confounding variable that impacts PD risk^77,78^. In our study, the association between PD severity and hysterectomies with the removal of ovaries indicates that this procedure is not only a risk factor but also an influence on the severity of the disease itself. It is critical for further studies to elucidate a definitive answer on the implications partial and total hysterectomies have in increasing the risk of PD and modulating PD severity, especially since they are prevalent procedures in gynecological care.

To our knowledge, this study is the first exploratory study that addresses sex-specific experiences from menses to menopause; however, there are some limitations. First, since this study primarily gathers WSHF history from a questionnaire, like other survey studies, it relies heavily on the response rate. Our response rate is low at 31.5%, while 60-80% is generally expected^79,80^. Although we are below average, a further limitation of our questionnaire response rate is selection bias since our questionnaire was deployed via email to PD GENEration, leaving those who are neither able to join PD GENEration nor have regular email access unable to participate. As PD GENEration is rapidly growing, we hope to continue deploying this questionnaire to women with PD. We have also begun distributing this questionnaire across North America Cleveland Clinic campuses. Currently, this questionnaire has been circulated nationally in the United States, and we are expanding our efforts to Latin America through the Latin American Research Consortium (LARGE-PD)^81^. This international effort will help better understand WSHF differences between regions and sociocultural norms, resulting in a better discussion of women and PD. These greater efforts will hopefully overcome the low response rate (due to selection bias) currently present in this study.

Another limitation is that this study uses PD severity. PD severity varies across literature and clinical standpoints, as many factors contribute to categorizing PD patients’ current health-related quality of life. We implemented Martinez-Martin et al.’s thresholds for severity, but because our study uses only one UPDRS time-point for analysis, this could be a misrepresentation of the respondents’ PD severity phenotype and result in misclassification bias. The UPDRS assessment can be influenced by many factors, such as the time medication was taken before the assessment, stress, “on” and “off” states, and whether the participant or clinician answered the questions^82^. Further studies will be needed to determine whether these thresholds represent the moderate/severe PD phenotype and use multiple UPDRS assessments. Progression of PD is another critical factor in the health-related quality of life, and it varies from person to person. Thus, studying the effects of WSHFs in PD progression will significantly benefit women with PD and the scientific community.

Lastly, sample size is another limitation of this study. Due to the overall n= 304, we have small sample sizes between our phenotype groups when looking at each WSHF. A larger sample size would allow us, with greater statistical power, to look at smaller effect sizes each WSHF has with PD severity. With a higher response rate, we hope to overcome this limitation by validating our findings in a larger sample size of women with PD.

Despite these limitations, our study has considerable strengths. We created a scientific questionnaire that addresses multiple WSHFs, some of which have never been addressed before^18^. Our questionnaire was a collaboration between neurologists, PD experts, and women’s health specialists and was validated for comprehension at Cleveland Clinic. Unlike other previous questionnaires, this one is publicly available for the sole purpose of incorporating WSHF questions into routine PD assessments. As many questionnaires used in the clinic are focused on gender-neutral factors, we hope this study sheds light on the purpose of acknowledging sex-specific aspects in the pursuit of bettering the treatment and care for those suffering from PD.

Previous studies have considered estrogen as a protective factor against PD. Although this has not been reproducible across the field, there are limited studies that have addressed estrogen in PD severity^38,39,41,43,83^. While our cohort did not use either estrogen, progesterone, or a combination of both hormone therapies, we did not see any exogenous hormone replacement or usage of birth control being associated with the mild phenotype. Further studies are needed to better understand the role of estrogen and hormones in PD severity.

Another strength of this study is that it considers the interactions between genetics, demographics, and clinical history when looking at PD severity. Previous literature that looked at WSHFs used electronic medical records or a questionnaire that didn’t incorporate genetics, leading them to be neglected in PD etiology. We had a moderate cohort of those with *LRRK2* and GBA mutations. Our study showed that amongst our cohort, having a *LRRK2* genotype was significantly associated with a moderate/severe phenotype. Thus, including genetic data in these models elucidates that genetic predisposition may be related to having a more severe PD phenotype within women.

Overall, this study used a questionnaire deployed across the United States to determine what WSHFs women with PD experienced. With this questionnaire, we were able to create a dataset of 304 women and their genetics, women-specific experiences, and clinical history. We found that by creating score thresholds for each UPDRS subpart, we could determine if WSHFs were associated with PD severity when adjusted for age, age at diagnosis, and PD medication. We found that depression and vaginal birth (Part I); depression and perinatal depression (Part II); *LRRK2* mutation status, B12 deficiency, and total hysterectomy (Part III) were significantly associated with a moderate/severe PD phenotype compared to a mild phenotype. This is one of the few studies that emphasize the role of WSHFs in PD. Future studies with a larger population of diverse women are needed to validate these findings, preferably with multiple UPDRS scores and a more detailed clinical history. Nevertheless, this study sets the groundwork for acknowledging the role WSHFs may play in PD and the potential benefit the scientific community can gain for therapeutics and clinical guidance if we further investigate the role sex-specific factors have in PD etiology.

## Data Availability

All data produced in the present study are available upon reasonable request to the authors

## Author contributions

1. Research project: A. Conception, B. Organization, C. Execution; 2. Statistical Analysis: A. Design, B. Execution, C. Review and Critique; 3. Manuscript Preparation: A. Writing of the first draft, B. Review and Critique

SCR: 1A, 1B, 1C, 2B, 3A, 3B, 3C

YL and BL: 2A, 2B, 2C, 3B

SP, TPL, and AS: 3C

KG, AN, PAS, and NG: 1B, 1C, 3B

MDL and HF: 1B, 1C, 3B

IFM: 1A, 1B, 1C, 2B, 3A, 3B, 3C

## Ethical Compliance Statement

The authors confirm that the approval of an institutional review board. We confirm that we have read the Journal’s position on issues involved in ethical publication and affirm that this work is consistent with those guidelines.

## Acknowledgments

We would like to acknowledge the Parkinson’s Foundation for this study. I.FM. and S.C.R had full access to all the data in the study and take responsibility for the integrity of the data and the accuracy of the data analysis.

