## Supplemental Table 1 for "Association of Women-Specific Health Factors in the Severity of Parkinson’s Disease"

| **Supplemental Table 2A. Part I: Univariate Logistic Regression Results** | | | |
| --- | --- | --- | --- |
|  | **Sample Size** | **OR (95% CI)** | **p-value** |
| **Age** |  |  |  |
|  | 160 | 1.01 (0.97, 1.05) | 0.6 |
| ***Hispanic*** |  |  |  |
|  | 160 | 0.32 (0.01, 10.05) | 0.52 |
| **White** |  |  |  |
|  | 160 | 2.20 (0.25, 19.32) | 0.48 |
| **Genetic Status** | 158 |  |  |
| GBA |  | 0.31 (0.05, 1.83) | 0.2 |
| LRRK2 |  | 0.63 (0.11, 3.71) | 0.61 |
| GBA | 160 | 0.69 (0.21, 2.24) | 0.54 |
| LRRK2 | 160 | 1.51 (0.47, 4.88) | 0.49 |
| PRKN | 160 | 5.00 (0.88, 28.29) | 0.069 |
| None |  | 0.41 (0.10, 1.74) | 0.23 |
| Other |  | -reference- |  |
| **Medications** |  |  |  |
| Levodopa | 160 | 1.66 (0.74, 3.70) | 0.22 |
| *Entacapone* | 159 | 17.14 (0.55, 533.12) | 0.11 |
| Carbidopa, Levodopa, and Entacapone (Stalevo) | 158 | 2.56 (0.78, 8.40) | 0.12 |
| Pramipexole | 158 | 1.42 (0.55, 3.66) | 0.47 |
| Ropinirole | 158 | 4.68 (1.30, 16.85) | ***0.018*** |
| Rotigotine | 158 | 1.19 (0.21, 6.72) | 0.84 |
| Rasagiline | 158 | 0.71 (0.29, 1.70) | 0.44 |
| Selegiline (oral, sublingual) | 159 | 1.16 (0.10, 13.10) | 0.9 |
| Amantadine (liquid, infusion) | 158 | 0.47 (0.13, 1.73) | 0.26 |
| **Current Comorbidities** |  |  |  |
| Arrythmia/Atrial Fibrillation | 160 | 1.00 (0.25, 4.04) | 0.99 |
| Arthritis | 160 | 1.96 (0.97, 3.94) | 0.059 |
| B12 deficiency | 160 | 1.95 (0.68, 5.60) | 0.21 |
| Cancer | 160 | 1.17 (0.21, 6.64) | 0.86 |
| Depression | 160 | 6.17 (2.89, 13.19) | ***<0.001*** |
| Diabetes Mellitus (adult onset) | 160 | 2.42 (0.47, 12.45) | 0.29 |
| Hearing Loss | 160 | 1.94 (0.91, 4.12) | 0.087 |
| Hyper cholesterolemia (High Cholesterol) | 160 | 1.50 (0.72, 3.13) | 0.28 |
| Hypertension (High Blood Pressure) | 160 | 0.82 (0.34, 1.99) | 0.66 |
| Loss of smell | 160 | 0.84 (0.42, 1.68) | 0.63 |
| Lung disease (including emphysema) | 160 | 2.05 (0.60, 7.09) | 0.25 |
| Other | 160 | 2.11 (1.06, 4.21) | ***0.034*** |
| Peripheral neuropathy | 160 | 0.69 (0.21, 2.24) | 0.54 |
| PTSD | 160 | 5.19 (1.24, 21.71) | ***0.024*** |
| Recreational drug use | 160 | 1.18 (0.28, 4.92) | 0.82 |
| Renal insufficiency (kidney disease) | 160 | 2.39 (0.33, 17.49) | 0.39 |
| Thyroid disease (not cancer, including Grave's disease) | 160 | 1.53 (0.68, 3.46) | 0.3 |
| **Still Menstruating Variable** |  |  |  |
|  | 160 | 1.07 (0.35, 3.26) | 0.91 |
| **Diagnosis of PMS Variable** |  |  |  |
|  | 160 | 1.86 (0.61, 5.68) | 0.28 |
| **Currently Perimenopausal Variable** |  |  |  |
|  | 156 | 0.96 (0.24, 3.89) | 0.96 |
| ***Hormone Levels Checked for Perimenopausal Variable*** |  |  |  |
|  | 10 | 15.40 (0.41, 584.08) | 0.14 |
| **Have Experienced Menopause Variable** |  |  |  |
|  | 157 | 1.21 (0.44, 3.28) | 0.71 |
| **Menses and PD Medication** |  |  |  |
| Medication felt less effective | 160 | 10.09 (1.10, 92.81) | ***0.041*** |
| More off/irregular frequency periods | 160 | 5.00 (0.88, 28.29) | 0.069 |
| **Hormone Replacement Therapy Variable** |  |  |  |
|  | 152 | 1.48 (0.69, 3.15) | 0.31 |
| **Birth Control Variable** |  |  |  |
|  | 110 | 0.68 (0.07, 6.74) | 0.74 |
| **Experience of Breast Feeding** | 159 |  |  |
| Yes |  | 1.62 (0.64, 4.12) | 0.31 |
| No |  | 1.60 (0.47, 5.47) | 0.45 |
| N/A |  | -reference- |  |
| **PD Onset** | 157 |  |  |
| While I was still having regular periods |  | -reference- |  |
| While I was going through perimenopause |  | 1.58 (0.42, 5.94) | 0.5 |
| One year or more after my last menstrual period |  | 1.00 (0.40, 2.50) | 0.99 |
| **History of Surgeries** |  |  |  |
| Total hysterectomy (removal of uterus, cervix, ovaries, Fallopian tubes, and surrounding structures) |  | 1.58 (0.66, 3.79) | 0.31 |
|  | 160 |  |  |
| Oophorectomy (surgical removal of one/unilateral or both/bilateral ovaries) | 160 | 1.86 (0.61, 5.68) | 0.28 |
| Mastectomy | 160 | 0.46 (0.05, 4.00) | 0.48 |
| Partial hysterectomy (just uterus) | 160 | 1.51 (0.47, 4.88) | 0.49 |
| **Types of Hormone Replacement Therapy** |  |  |  |
| Estrogen | 160 | 1.74 (0.62, 4.89) | 0.29 |
| Progesterone/Progestin | 160 | 1.58 (0.26, 9.77) | 0.62 |
| Estrogen and progesterone combination | 160 | 1.00 (0.36, 2.78) | 0.99 |
| **Types of Birth Control** |  |  |  |
| Pill | 160 | 1.66 (0.76, 3.62) | 0.2 |
| Ring | 160 | 0.57 (0.06, 5.28) | 0.62 |
| Hormonal IUD | 160 | 0.76 (0.20, 2.95) | 0.7 |
| Copper IUD | 160 | 1.46 (0.50, 4.27) | 0.49 |
| Tubal ligation | 160 | 4.33 (1.64, 11.45) | ***0.003*** |
| **Pregnancy Experiences** |  |  |  |
| *In vitro fertilization* | 160 | 0.25 (0.01, 6.63) | 0.41 |
| C-section | 160 | 0.55 (0.21, 1.46) | 0.23 |
| Vaginal birth | 160 | 3.48 (1.49, 8.13) | ***0.004*** |
| Having children prior to PD diagnosis | 160 | 3.44 (1.25, 9.47) | ***0.017*** |
| No children | 160 | 0.35 (0.11, 1.08) | 0.068 |
| **Difficulties in Pregnancy Experiences** |  |  |  |
| Conceiving (fertility) | 160 | 1.76 (0.70, 4.44) | 0.23 |
| Childbirth | 160 | 3.03 (1.19, 7.73) | ***0.02*** |
| Pregnancy (reaching full birth) | 160 | 1.40 (0.55, 3.60) | 0.48 |
| **Pregnancy-related Aliments** |  |  |  |
| Eclampsia or pre-eclampsia | 160 | 0.57 (0.12, 2.77) | 0.48 |
| *Diabetes during pregnancy* | 160 | 28.47 (1.17, 692.41) | ***0.04*** |
| *Viral infection* | 160 | 7.02 (0.08, 647.52) | 0.4 |
| Bacterial infection | 160 | 5.00 (0.88, 28.29) | 0.069 |
| Depression/anxiety during pregnancy | 160 | 22.20 (2.69, 183.12) | ***0.004*** |
| Postpartum Depression | 160 | 6.36 (2.07, 19.52) | ***0.001*** |
| Flair up migraines | 160 | 3.02 (0.96, 9.51) | 0.06 |
| **Hormone-related Disorders** |  |  |  |
| Polycystic Ovarian Syndrome | 160 | 3.30 (0.71, 15.37) | 0.13 |
| Hypothyroidism | 160 | 1.12 (0.45, 2.80) | 0.81 |
| Hyperthyroidism | 160 | 0.57 (0.12, 2.77) | 0.48 |
| Diabetes Type II | 160 | 0.93 (0.17, 4.97) | 0.93 |
| Migraines (no aura) | 160 | 2.12 (0.82, 5.51) | 0.12 |

*Variables in italics:* use Firth method

 Dummy variables for each response option

Reference was used to for univariate modeling and for the outcome of moderate/severe phenotype

| **Supplemental Table 2B. Part II: Univariate Logistic Regression Results** | | | |
| --- | --- | --- | --- |
|  | **Sample Size** | **OR (95% CI)** | **p-value** |
| **Age** |  |  |  |
|  | 302 | 1.02 (0.98, 1.05) | 0.33 |
| **Hispanic** |  |  |  |
|  | 300 | 0.72 (0.09, 6.11) | 0.76 |
| ***White*** |  |  |  |
|  | 299 | 6.62 (0.35, 125.61) | 0.21 |
| **Genetic Status** | 299 |  |  |
| GBA |  | 1.07 (0.26, 4.31) | 0.93 |
| LRRK2 |  | 0.50 (0.10, 2.58) | 0.41 |
| GBA | 302 | 1.39 (0.56, 3.41) | 0.48 |
| LRRK2 | 302 | 0.61 (0.17, 2.11) | 0.43 |
| PRKN | 302 | 1.68 (0.43, 6.56) | 0.45 |
| None |  | 0.80 (0.25, 2.54) | 0.7 |
| Other |  | -reference- |  |
| **Medications** |  |  |  |
| Levodopa | 302 | 2.20 (1.03, 4.73) | ***0.043*** |
| Entacapone | 300 | 1.77 (0.33, 9.37) | 0.5 |
| Carbidopa, Levodopa, and Entacapone (Stalevo) | 299 | 1.96 (0.72, 5.36) | 0.19 |
| Pramipexole | 299 | 1.30 (0.53, 3.20) | 0.56 |
| Ropinirole | 299 | 2.72 (1.13, 6.52) | ***0.025*** |
| Rotigotine | 299 | 3.31 (1.01, 10.83) | ***0.048*** |
| Rasagiline | 299 | 0.52 (0.23, 1.16) | 0.11 |
| Selegiline (oral, sublingual) | 300 | 0.30 (0.04, 2.32) | 0.25 |
| Amantadine (liquid, infusion) | 299 | 2.09 (0.99, 4.43) | 0.053 |
| **Current Comorbidities** |  |  |  |
| Arrythmia/Atrial Fibrillation | 302 | 1.24 (0.44, 3.50) | 0.68 |
| Arthritis | 302 | 1.06 (0.59, 1.89) | 0.85 |
| B12 deficiency | 302 | 1.00 (0.36, 2.76) | 0.99 |
| Cancer | 302 | 1.62 (0.56, 4.71) | 0.37 |
| Depression | 302 | 2.94 (1.61, 5.38) | ***<0.001*** |
| Diabetes Mellitus (adult onset) | 302 | 0.87 (0.19, 4.10) | 0.86 |
| Hearing Loss | 302 | 1.76 (0.95, 3.27) | 0.074 |
| Hyper cholesterolemia (High Cholesterol) | 302 | 0.63 (0.32, 1.27) | 0.2 |
| Hypertension (High Blood Pressure) | 302 | 1.27 (0.65, 2.47) | 0.48 |
| Loss of smell | 302 | 1.93 (1.08, 3.47) | ***0.027*** |
| Lung disease (including emphysema) | 302 | 1.51 (0.53, 4.34) | 0.44 |
| Other | 302 | 0.97 (0.53, 1.79) | 0.93 |
| Peripheral neuropathy | 302 | 2.93 (1.38, 6.23) | ***0.005*** |
| PTSD | 302 | 1.50 (0.47, 4.84) | 0.5 |
| Recreational drug use | 302 | 0.66 (0.15, 3.03) | 0.6 |
| Renal insufficiency (kidney disease) | 302 | 3.43 (0.74, 15.76) | 0.11 |
| Thyroid disease (not cancer, including Grave's disease) | 302 | 1.25 (0.62, 2.50) | 0.53 |
| **Still Menstruating Variable** |  |  |  |
|  | 301 | 0.17 (0.02, 1.26) | 0.083 |
| **Diagnosis of PMS Variable** |  |  |  |
|  | 301 | 1.20 (0.49, 2.93) | 0.68 |
| **Currently Perimenopausal Variable** |  |  |  |
|  | 294 | 0.53 (0.12, 2.36) | 0.4 |
| ***Hormone Levels Checked for Perimenopausal Variable*** |  |  |  |
|  | 18 | 1.29 (0.07, 24.38) | 0.87 |
| **Have Experienced Menopause Variable** |  |  |  |
|  | 298 | 9.09 (1.22, 67.83) | ***0.031*** |
| **Menses and PD Medication** |  |  |  |
| Medication felt less effective | 302 | 4.65 (1.13, 19.21) | ***0.034*** |
| More off/irregular frequency periods | 302 | 3.08 (0.84, 11.29) | 0.09 |
| **Hormone Replacement Therapy Variable** |  |  |  |
|  | 288 | 1.36 (0.73, 2.55) | 0.33 |
| **Birth Control Variable** |  |  |  |
|  | 202 | 0.55 (0.07, 4.50) | 0.57 |
| **Experience of Breast Feeding** | 296 |  |  |
| Yes |  | 0.86 (0.40, 1.84) | 0.7 |
| No |  | 1.26 (0.49, 3.23) | 0.63 |
| N/A |  | -reference- |  |
| **PD Onset** | 296 |  |  |
| While I was still having regular periods |  | -reference- |  |
| While I was going through perimenopause |  | 0.90 (0.30, 2.72) | 0.85 |
| One year or more after my last menstrual period |  | 0.59 (0.29, 1.22) | 0.16 |
| **History of Surgeries** |  |  |  |
| Total hysterectomy (removal of uterus, cervix, ovaries, Fallopian tubes, and surrounding structures) | 302 | 1.59 (0.77, 3.30) | 0.21 |
| Oophorectomy (surgical removal of one/unilateral or both/bilateral ovaries) | 302 | 0.58 (0.17, 2.00) | 0.38 |
| Mastectomy | 302 | 5.93 (1.54, 22.85) | ***0.01*** |
| Partial hysterectomy (just uterus) | 302 | 0.58 (0.19, 1.71) | 0.32 |
| **Types of Hormone Replacement Therapy** |  |  |  |
| Estrogen | 302 | 1.25 (0.54, 2.90) | 0.61 |
| Progesterone/Progestin | 302 | 1.10 (0.23, 5.34) | 0.9 |
| Estrogen and progesterone combination | 302 | 1.98 (0.92, 4.29) | 0.082 |
| **Types of Birth Control** |  |  |  |
| Pill | 302 | 0.89 (0.48, 1.64) | 0.7 |
| Ring | 302 | 1.48 (0.29, 7.54) | 0.64 |
| Hormonal IUD | 302 | 1.27 (0.40, 4.03) | 0.68 |
| Copper IUD | 302 | 0.73 (0.27, 1.99) | 0.54 |
| Tubal ligation | 302 | 1.11 (0.46, 2.69) | 0.81 |
| **Pregnancy Experiences** |  |  |  |
| *In vitro fertilization* | 302 | 0.33 (0.01, 7.42) | 0.48 |
| C-section | 302 | 1.01 (0.49, 2.09) | 0.98 |
| Vaginal birth | 302 | 1.13 (0.62, 2.07) | 0.69 |
| Having children prior to PD diagnosis | 302 | 0.96 (0.50, 1.85) | 0.91 |
| No children | 302 | 0.93 (0.45, 1.94) | 0.86 |
| **Difficulties in Pregnancy Experiences** |  |  |  |
| Conceiving (fertility) | 302 | 0.89 (0.37, 2.13) | 0.79 |
| Childbirth | 302 | 1.16 (0.48, 2.81) | 0.74 |
| Pregnancy (reaching full birth) | 302 | 1.06 (0.41, 2.72) | 0.9 |
| **Pregnancy-related Aliments** |  |  |  |
| Eclampsia or pre-eclampsia | 302 | 0.30 (0.04, 2.34) | 0.25 |
| Diabetes during pregnancy | 302 | 0.73 (0.09, 6.16) | 0.77 |
| Viral infection | 302 | 2.24 (0.40, 12.55) | 0.36 |
| Bacterial infection | 302 | 2.63 (0.74, 9.30) | 0.13 |
| Depression/anxiety during pregnancy | 302 | 5.78 (1.70, 19.69) | ***0.005*** |
| Postpartum Depression | 302 | 2.11 (0.87, 5.14) | 0.1 |
| Flair up migraines | 302 | 2.56 (0.97, 6.75) | 0.057 |
| **Hormone-related Disorders** |  |  |  |
| Polycystic Ovarian Syndrome | 302 | 1.91 (0.65, 5.67) | 0.24 |
| Hypothyroidism | 302 | 1.00 (0.47, 2.13) | 0.99 |
| Hyperthyroidism | 302 | 0.81 (0.23, 2.89) | 0.75 |
| Diabetes Type II | 302 | 0.66 (0.15, 3.03) | 0.6 |
| Migraines (no aura) | 302 | 1.69 (0.77, 3.73) | 0.19 |

*Variables in italics:* use Firth method

 Dummy variables for each response option

Reference was used to for univariate modeling and for the outcome of moderate/severe phenotype

| **Supplemental Table 2C. Part III: Univariate Logistic Regression Results** | | | |
| --- | --- | --- | --- |
|  | **Sample Size** | **OR (95% CI)** | **p-value** |
| **Age** |  |  |  |
|  | 141 | 1.01 (0.96, 1.06) | 0.72 |
| ***Hispanic*** |  |  |  |
|  | 141 | 0.58 (0.02, 18.20) | 0.76 |
| **White** |  |  |  |
|  | 141 | 0.70 (0.07, 7.03) | 0.76 |
| **Genetic Status** | 139 |  |  |
| GBA |  | 1.20 (0.17, 8.66) | 0.86 |
| LRRK2 |  | 2.50 (0.34, 18.33) | 0.37 |
| GBA | 141 | 1.81 (0.52, 6.28) | 0.35 |
| LRRK2 | 141 | 4.09 (1.15, 14.60) | ***0.03*** |
| PRKN | 141 | 2.20 (0.38, 12.69) | 0.38 |
| None |  | 0.53 (0.10, 2.88) | 0.47 |
| Other |  | -reference- |  |
| **Medications** |  |  |  |
| Levodopa | 141 | 0.68 (0.28, 1.69) | 0.41 |
| Entacapone | 140 | 2.13 (0.19, 24.45) | 0.54 |
| Carbidopa, Levodopa, and Entacapone (Stalevo) | 139 | 1.04 (0.21, 5.20) | 0.96 |
| Pramipexole | 139 | 1.59 (0.52, 4.88) | 0.42 |
| Ropinirole | 139 | 0.44 (0.05, 3.63) | 0.45 |
| Rotigotine | 139 | 6.87 (1.09, 43.41) | ***0.04*** |
| Rasagiline | 139 | 0.83 (0.29, 2.43) | 0.74 |
| *Selegiline (oral, sublingual)* | 140 | 0.57 (0.02, 18.04) | 0.75 |
| Amantadine (liquid, infusion) | 139 | 1.77 (0.51, 6.16) | 0.37 |
| **Current Comorbidities** |  |  |  |
| Arrythmia/Atrial Fibrillation | 141 | 0.51 (0.06, 4.26) | 0.53 |
| Arthritis | 141 | 0.54 (0.23, 1.26) | 0.15 |
| B12 deficiency | 141 | 4.64 (1.51, 14.23) | ***0.007*** |
| Cancer | 141 | 1.06 (0.11, 9.87) | 0.96 |
| Depression | 141 | 1.40 (0.57, 3.45) | 0.46 |
| Diabetes Mellitus (adult onset) | 141 | 2.20 (0.38, 12.69) | 0.38 |
| Hearing Loss | 141 | 0.84 (0.31, 2.28) | 0.73 |
| Hyper cholesterolemia (High Cholesterol) | 141 | 0.90 (0.35, 2.33) | 0.82 |
| Hypertension (High Blood Pressure) | 141 | 1.27 (0.45, 3.52) | 0.65 |
| Loss of smell | 141 | 1.65 (0.71, 3.84) | 0.24 |
| Lung disease (including emphysema) | 141 | 2.25 (0.53, 9.64) | 0.27 |
| Other | 141 | 0.41 (0.15, 1.09) | 0.073 |
| Peripheral neuropathy | 141 | 0.75 (0.16, 3.60) | 0.72 |
| PTSD | 141 | 2.73 (0.61, 12.19) | 0.19 |
| Recreational drug use | 141 | 0.59 (0.07, 4.99) | 0.63 |
| *Renal insufficiency (kidney disease)* | 141 | 0.58 (0.02, 18.20) | 0.76 |
| Thyroid disease (not cancer, including Grave's disease) | 141 | 0.65 (0.21, 2.07) | 0.47 |
| **Still Menstruating Variable** |  |  |  |
|  | 141 | 1.06 (0.28, 4.06) | 0.93 |
| **Diagnosis of PMS Variable** |  |  |  |
|  | 141 | 1.46 (0.37, 5.80) | 0.59 |
| ***Currently Perimenopausal Variable*** |  |  |  |
|  | 137 | 0.18 (0.01, 3.69) | 0.27 |
| ***Hormone Levels Checked for Perimenopausal Variable*** |  |  |  |
| **Have Experienced Menopause Variable** |  |  |  |
|  | 138 | 0.70 (0.23, 2.12) | 0.53 |
| **Menses and PD Medication** |  |  |  |
| Medication felt less effective | 141 | 2.15 (0.19, 24.66) | 0.54 |
| More off/irregular frequency periods | 141 | 1.06 (0.11, 9.87) | 0.96 |
| **Hormone Replacement Therapy Variable** |  |  |  |
|  | 133 | 1.97 (0.81, 4.79) | 0.13 |
| ***Birth Control Variable*** |  |  |  |
|  | 100 | 0.41 (0.02, 11.28) | 0.6 |
| **Experience of Breast Feeding** | 140 |  |  |
| Yes |  | 3.33 (0.73, 15.28) | 0.12 |
| No |  | 4.81 (0.82, 28.27) | 0.082 |
| N/A |  | -reference- |  |
| **PD Onset** | 138 |  |  |
| While I was still having regular periods |  | -reference- |  |
| While I was going through perimenopause |  | 3.43 (0.79, 14.85) | 0.1 |
| One year or more after my last menstrual period |  | 0.76 (0.25, 2.33) | 0.63 |
| **History of Surgeries** |  |  |  |
| Total hysterectomy (removal of uterus, cervix, ovaries, Fallopian tubes, and surrounding structures) | 141 | 3.30 (1.25, 8.68) | ***0.016*** |
| Oophorectomy (surgical removal of one/unilateral or both/bilateral ovaries) | 141 | 0.36 (0.04, 2.92) | 0.34 |
| Mastectomy | 141 | 1.06 (0.11, 9.87) | 0.96 |
| *Partial hysterectomy (just uterus)* | 141 | 0.15 (0.01, 2.91) | 0.21 |
| **Types of Hormone Replacement Therapy** |  |  |  |
| Estrogen | 141 | 2.13 (0.67, 6.74) | 0.2 |
| *Progesterone/Progestin* | 141 | 0.36 (0.02, 8.87) | 0.53 |
| Estrogen and progesterone combination | 141 | 2.43 (0.82, 7.20) | 0.11 |
| **Types of Birth Control** |  |  |  |
| Pill | 141 | 1.11 (0.43, 2.89) | 0.82 |
| Ring | 141 | 1.42 (0.14, 14.24) | 0.76 |
| Hormonal IUD | 141 | 1.66 (0.41, 6.71) | 0.48 |
| Copper IUD | 141 | 0.68 (0.14, 3.24) | 0.63 |
| Tubal ligation | 141 | 0.97 (0.26, 3.68) | 0.97 |
| **Pregnancy Experiences** |  |  |  |
| *In vitro fertilization* | 141 | 0.45 (0.02, 12.03) | 0.63 |
| C-section | 141 | 1.34 (0.48, 3.75) | 0.57 |
| Vaginal birth | 141 | 1.04 (0.43, 2.53) | 0.93 |
| Having children prior to PD diagnosis | 141 | 1.30 (0.45, 3.77) | 0.63 |
| No children | 141 | 0.59 (0.16, 2.14) | 0.42 |
| **Difficulties in Pregnancy Experiences** |  |  |  |
| Conceiving (fertility) | 141 | 0.63 (0.17, 2.29) | 0.48 |
| Childbirth | 141 | 0.46 (0.10, 2.11) | 0.31 |
| Pregnancy (reaching full birth) | 141 | 1.50 (0.49, 4.56) | 0.47 |
| **Pregnancy-related Aliments** |  |  |  |
| Eclampsia or pre-eclampsia | 141 | 3.79 (0.94, 15.22) | 0.06 |
| Diabetes during pregnancy | 141 | 1.42 (0.14, 14.24) | 0.76 |
| Bacterial infection | 141 | 9.04 (0.79, 103.63) | 0.077 |
| Depression/anxiety during pregnancy | 141 | 0.69 (0.08, 6.00) | 0.74 |
| *Postpartum Depression* | 141 | 0.13 (0.01, 2.41) | 0.17 |
| Flair up migraines | 141 | 0.93 (0.19, 4.59) | 0.93 |
| **Hormone-related Disorders** |  |  |  |
| Polycystic Ovarian Syndrome | 141 | 0.84 (0.09, 7.49) | 0.87 |
| Hypothyroidism | 141 | 0.63 (0.17, 2.29) | 0.48 |
| *Hyperthyroidism* | 141 | 0.20 (0.01, 4.17) | 0.3 |
| Diabetes Type II | 141 | 2.20 (0.38, 12.69) | 0.38 |
| Migraines (no aura) | 141 | 2.68 (0.89, 8.04) | 0.079 |

*Variables in italics:* use Firth method

 Dummy variables for each response option

Reference was used to for univariate modeling and for the outcome of moderate/severe phenotype

| **Supplemental Table 2D. Part IV: Univariate Logistic Regression Results** | | | | |
| --- | --- | --- | --- | --- |
|  | **Sample Size** | | **OR (95% CI)** | **p-value** |
| **Age** | |  |  |  |
|  | | 139 | 1.01 (0.97, 1.05) | 0.64 |
| ***Hispanic*** | |  |  |  |
|  | | 139 | 0.31 (0.01, 9.58) | 0.5 |
| **White** | |  |  |  |
|  | | 139 | 2.31 (0.26, 20.37) | 0.45 |
| **Genetic Status** | | 138 |  |  |
| GBA | |  | 2.63 (0.40, 17.46) | 0.32 |
| LRRK2 | |  | 3.00 (0.40, 22.71) | 0.29 |
| GBA | | 139 | 2.14 (0.72, 6.34) | 0.17 |
| LRRK2 | | 139 | 2.39 (0.66, 8.75) | 0.19 |
| PRKN | | 139 | 1.12 (0.20, 6.37) | 0.9 |
| None | |  | 1.15 (0.22, 6.00) | 0.87 |
| Other | |  | -reference- |  |
| **Medications** | |  |  |  |
| Levodopa | | 139 | 1.42 (0.52, 3.88) | 0.49 |
| Entacapone | | 138 | 1.11 (0.10, 12.55) | 0.93 |
| Carbidopa, Levodopa, and Entacapone (Stalevo) | | 137 | 2.38 (0.72, 7.86) | 0.16 |
| Pramipexole | | 137 | 1.11 (0.41, 2.99) | 0.83 |
| Ropinirole | | 137 | 1.93 (0.56, 6.71) | 0.3 |
| Rotigotine | | 137 | 4.72 (0.83, 26.83) | 0.08 |
| Rasagiline | | 137 | 0.82 (0.34, 1.95) | 0.65 |
| Selegiline (oral, sublingual) | | 138 | 1.11 (0.10, 12.55) | 0.93 |
| Amantadine (liquid, infusion) | | 137 | 2.46 (0.85, 7.06) | 0.095 |
| **Current Comorbidities** | |  |  |  |
| Arrythmia/Atrial Fibrillation | | 139 | 0.89 (0.17, 4.77) | 0.89 |
| Arthritis | | 139 | 0.93 (0.45, 1.92) | 0.85 |
| B12 deficiency | | 139 | 2.91 (0.98, 8.62) | 0.054 |
| Cancer | | 139 | 0.43 (0.05, 3.83) | 0.45 |
| Depression | | 139 | 2.42 (1.12, 5.23) | ***0.024*** |
| Diabetes Mellitus (adult onset) | | 139 | 0.43 (0.05, 3.83) | 0.45 |
| Hearing Loss | | 139 | 1.02 (0.43, 2.39) | 0.96 |
| Hyper cholesterolemia (High Cholesterol) | | 139 | 0.79 (0.35, 1.79) | 0.58 |
| Hypertension (High Blood Pressure) | | 139 | 0.87 (0.35, 2.16) | 0.76 |
| Loss of smell | | 139 | 2.21 (1.07, 4.61) | ***0.033*** |
| Lung disease (including emphysema) | | 139 | 1.97 (0.57, 6.86) | 0.28 |
| Other | | 139 | 1.25 (0.59, 2.62) | 0.56 |
| Peripheral neuropathy | | 139 | 1.02 (0.33, 3.13) | 0.98 |
| PTSD | | 139 | 4.08 (0.93, 17.92) | 0.063 |
| Recreational drug use | | 139 | 0.62 (0.12, 3.12) | 0.56 |
| Renal insufficiency (kidney disease) | | 139 | 2.29 (0.31, 16.84) | 0.41 |
| Thyroid disease (not cancer, including Grave's disease) | | 139 | 0.95 (0.39, 2.28) | 0.9 |
| **Still Menstruating Variable** | |  |  |  |
|  | | 139 | 0.18 (0.02, 1.47) | 0.11 |
| **Diagnosis of PMS Variable** | |  |  |  |
|  | | 139 | 2.43 (0.74, 8.03) | 0.14 |
| ***Currently Perimenopausal Variable*** | |  |  |  |
|  | | 136 | 0.62 (0.12, 3.13) | 0.56 |
| ***Hormone Levels Checked for Perimenopausal Variable*** | |  |  |  |
|  | | 9 | 0.75 (0.03, 17.51) | 0.86 |
| **Have Experienced Menopause Variable** | |  |  |  |
|  | | 136 | 1.52 (0.47, 4.99) | 0.49 |
| **Menses and PD Medication** | |  |  |  |
| Medication felt less effective | | 139 | 9.74 (1.06, 89.96) | ***0.045*** |
| More off/irregular frequency periods | | 139 | 4.82 (0.85, 27.41) | 0.076 |
| **Hormone Replacement Therapy Variable** | | 132 | 0.57 (0.24, 1.35) | 0.2 |
| **Birth Control Variable** | |  |  |  |
|  | | 96 | 0.69 (0.07, 6.90) | 0.75 |
| **Experience of Breast Feeding** | | 138 |  |  |
| Yes | |  | 1.36 (0.49, 3.78) | 0.55 |
| No | |  | 2.59 (0.73, 9.25) | 0.14 |
| N/A | |  | -reference- |  |
| **PD Onset** | | 136 |  |  |
| While I was still having regular periods | |  | -reference- |  |
| While I was going through perimenopause | |  | 1.94 (0.50, 7.64) | 0.34 |
| One year or more after my last menstrual period | |  | 0.63 (0.25, 1.60) | 0.33 |
| **History of Surgeries** | |  |  |  |
| Total hysterectomy (removal of uterus, cervix, ovaries, Fallopian tubes, and surrounding structures) | | 139 | 0.97 (0.37, 2.57) | 0.95 |
| Oophorectomy (surgical removal of one/unilateral or both/bilateral ovaries) | | 139 | 2.43 (0.74, 8.03) | 0.14 |
| Mastectomy | | 139 | 3.52 (0.57, 21.91) | 0.18 |
| Partial hysterectomy (just uterus) | | 139 | 0.99 (0.29, 3.42) | 0.99 |
| **Types of Hormone Replacement Therapy** | |  |  |  |
| Estrogen | | 139 | 1.02 (0.33, 3.13) | 0.98 |
| *Progesterone/Progestin* | | 139 | 0.19 (0.01, 4.66) | 0.31 |
| Estrogen and progesterone combination | | 139 | 0.84 (0.28, 2.53) | 0.76 |
| **Types of Birth Control** | |  |  |  |
| Pill | | 139 | 1.17 (0.53, 2.60) | 0.69 |
| *Ring* | | 139 | 0.24 (0.01, 6.32) | 0.39 |
| Hormonal IUD | | 139 | 0.83 (0.21, 3.27) | 0.78 |
| Copper IUD | | 139 | 2.47 (0.81, 7.55) | 0.11 |
| Tubal ligation | | 139 | 1.77 (0.65, 4.76) | 0.26 |
| **Pregnancy Experiences** | |  |  |  |
| In vitro fertilization | | 139 | 1.12 (0.10, 12.68) | 0.93 |
| C-section | | 139 | 0.66 (0.24, 1.78) | 0.41 |
| Vaginal birth | | 139 | 1.35 (0.62, 2.98) | 0.45 |
| Having children prior to PD diagnosis | | 139 | 2.17 (0.82, 5.76) | 0.12 |
| No children | | 139 | 0.75 (0.27, 2.07) | 0.58 |
| **Difficulties in Pregnancy Experiences** | |  |  |  |
| Conceiving (fertility) | | 139 | 1.04 (0.37, 2.94) | 0.95 |
| Childbirth | | 139 | 1.04 (0.37, 2.94) | 0.95 |
| Pregnancy (reaching full birth) | | 139 | 1.60 (0.60, 4.25) | 0.35 |
| **Pregnancy-related Aliments** | |  |  |  |
| Eclampsia or pre-eclampsia | | 139 | 3.18 (0.68, 14.87) | 0.14 |
| Diabetes during pregnancy | | 139 | 9.74 (1.06, 89.96) | ***0.045*** |
| *Viral infection* | | 139 | 6.72 (0.07, 620.49) | 0.41 |
| Bacterial infection | | 139 | 12.49 (1.41, 110.45) | ***0.023*** |
| Depression/anxiety during pregnancy | | 139 | 1.73 (0.37, 8.07) | 0.49 |
| Postpartum Depression | | 139 | 0.58 (0.15, 2.19) | 0.42 |
| Flair up migraines | | 139 | 5.03 (1.19, 21.16) | ***0.028*** |
| **Hormone-related Disorders** | |  |  |  |
| Polycystic Ovarian Syndrome | | 139 | 1.73 (0.37, 8.07) | 0.49 |
| Hypothyroidism | | 139 | 0.90 (0.34, 2.37) | 0.84 |
| Hyperthyroidism | | 139 | 1.13 (0.27, 4.72) | 0.87 |
| *Diabetes Type II* | | 139 | 0.16 (0.01, 3.65) | 0.25 |
| Migraines (no aura)  *Variables in italics:* use Firth method   Dummy variables for each response option  Reference was used to for univariate modeling and for the outcome of moderate/severe phenotype | | 139 | 0.84 (0.28, 2.53) | 0.76 |
