## Supplemental Table 2 for "Association of Women-Specific Health Factors in the Severity of Parkinson’s Disease"

### **Supplemental Table 1. Frequency of UPDRS Scores, by Severity Groups Parts I-IV**

|  | Part I | Part II | Part III | Part IV |
| --- | --- | --- | --- | --- |
| UPDRS Moderate/Severe Threshold | >21 | >29 | >58 | >12 |
| Mild n (%) | 112 (70.0) | 246 (81.46) | 114 (80.85) | 96 (69.06) |
| Moderate/Severe n (%) | 48 (30.0) | 56 (18.54) | 27 (19.15) | 40 (30.94) |
